# Exploring the exposome and unexplained variance in biological ageing – insights from a longitudinal twin study in adolescence and early adulthood

**DOI:** 10.64898/2026.03.03.26347499

**Authors:** A. Opperbeck, Z. Wang, I. Rautiainen, A. Heikkinen, J. Kaprio, M. Ollikainen, S. Sebert, E. Sillanpää

## Abstract

Biological ageing begins before birth, with early-life exposures shaping late-life health. These exposures drive health inequities early, yet specific exposures and the composition of the ageing exposome remain largely undefined. This gap may persist as the field lacks agnostic investigations accounting for non-linearity, interactions and subtle signals.

We aimed to identify exposures predictive of epigenetic ageing accumulated during childhood and adolescence and explore the composition of the “missing” exposome. In the FinnTwin12 cohort (847 participants measured at ages 12, 14, 17, and 22), over 500 exposures (including lifestyle, green environments, air pollutants, and demographic factors) were analysed using exposome-wide association studies and data-driven ML models (Knockoff Boosted Tree, sNPLS and Boruta). Epigenetic age (blood DNA methylation at age 22) was estimated using GrimAge and DunedinPACE. Our exposure set explains ∼28% of the variance in epigenetic age (R^2^ _GrimAge_ = 25.7%; R^2^ _DunedinPACE_ = 30.8%). Predictors of increased epigenetic age included lifestyle and socioeconomic factors (*smoking*, *alcohol use*, *youth unemployment*), alongside *green space*, while *tree cover*, *vegetation index*, *neighbourhood age structure* and *aerial black carbon* emerged as predictors of decreased epigenetic age. Twin modelling revealed that unexplained variance – the ‘missing exposome’ – consists primarily of environmental factors unshared by twin siblings, distinct from the substantial genetic component captured by our model.

Our results underscore the need to expand the exposome approach and model non-linearities to reveal subtle environmental signals accumulating early in life. Because identified predictors include modifiable systemic factors, they offer opportunities to alter health trajectories and mitigate inequity early on.

## 1. Introduction

Ageing is not a disease but a complex process increasing the likelihood of disease later in life. The biological foundations for late-life health are established at a young age. Ageing processes begin before birth and follow developmental plasticity, where environmental exposures in childhood influence adult traits and, later, susceptibility to disease via epigenomic alterations (Skinner, 2024). Many of the external drivers of ageing, and their interplay and long-term effects, are still not well understood because quantifying the progress of biological ageing pace in living humans is challenging.

Epigenetic age (EA), estimated through DNA methylation (DNAm) at cytosine-phosphate-guanine (CpG) sites, has been proposed as a hallmark of ageing and change of it, indicating either alignment with chronological age or acceleration/deceleration relative to chronological age. Methylation at specific CpG sites is regarded as indicative of intrinsic ageing processes as well as environmental exposure. Its subsequent altering of gene expression can be adaptive or maladaptive and thus e.g. forecasting future health (Colwell et al., 2023; Peters et al., 2021). Epigenetic clocks, i.e. algorithms trained on DNAm profiles, estimate EA, and newer clocks have evolved from predicting chronological age to projecting mortality (GrimAge (Lu et al., 2019) and morbidity (DunedinPACE (Belsky et al., 2022). They correlate with diverse outcomes, including frailty, cognitive decline, cardiovascular disease, diabetes, and metabolic syndrome (Faul et al., 2023; Föhr et al., 2024; Fraszczyk et al., 2022; Lu et al., 2019; McCrory et al., 2020; Oblak et al., 2021; Savin et al., 2024). Evidence suggests that early childhood is a critical period where environmental exposures can influence epigenetic ageing trajectories with long-term health consequences (Wang et al., 2023).

Among others, BMI, smoking history, alcohol use, physical activity, diet, mental health, socioeconomic status (SES), and education have been identified as drivers of EA. Most epidemiological studies have focused on single exposures or domains. While understanding the impact of specific exposures is fundamental to fathom disease aetiology, the single-exposure approach neglects the complexity of influences and the possibility of co-exposure confounding (Vrijheid, 2014). The human exposome concept responds to this omission. It is defined as the complete set of environmental and biological exposures that influence an individual’s health from conception throughout their life course (Miller & Jones, 2014; Wild, 2005). The concept distinguishes between general and specific external, and internal exposures. Understanding early life ageing and promoting health equity requires identifying these exposures, often indiscernible as they leave their imprint on our DNA (epigenetic modification) and consequent cell function.

Considering the presence of latent factors that often remain unrecognized, achieving exhaustive exposure sets is inherently difficult. Exposome research therefore seeks to capture the broadest possible range of exposures across multiple domains and examine them as multifactorial determinants of outcomes. Data-driven approaches can reveal previously unknown effects. While the current era offers unprecedented access to abundant exposome data, their heterogeneity, multicollinearity and dynamic nature across time and space render them challenging to study (Sainani, 2016). Despite the fact that early life exposures are known to significantly program later-life health, studies linking a comprehensive external exposome to epigenetic ageing have been limited by scope and/or methodology and often neglected the non-linear relationships and interactions characteristic of natural living systems. Exposome-wide association studies (ExWASs) in the HELIX cohort (Human Early Life Exposome) linked indoor fine particulate matter (PM_abs_) and passive smoking to EA (de Prado-Bert et al., 2021), and a wide range of environmental exposures including passive smoking, air quality, diet, weather and chemical pollutants correlated with changes in DNAm (Maitre et al., 2022). However, a recent HELIX/ATHLETE study (Amine et al., 2025) using more sophisticated correlation analyses could not find associations between either environmental exposures or a health score and DNAm. To date, we are not aware of an exposome study on EA in young adults that would also account for exposure interactions or non-linearity.

In this longitudinal twin study, we explore a large set of external exposures accumulated during childhood and adolescence for associations with EA in early adulthood by applying a systematic analysis pipeline. Our use of the twin design allows control of genetic confounding, strengthening the tests of environmental associations, and allows us to quantify the proportion of variance explained by our exposome dataset and provide novel information on the decomposition of variance left unexplained.

## 2. Methods

### 2.1 Study Population

This study was conducted using the FinnTwin12 (FT12) cohort. In brief, FT12 is a nation-wide prospective cohort of mono-and dizygotic (MZ, DZ) twins born between 1983 and 1987 (Kaprio, 2013; Rose et al., 2019a). A total of 5184 individuals (2705 families; 94% response rate) retrieved from the population register replied to the initial questionnaire (wave 1 at age 12) and entered the study upon satisfaction of inclusion criteria, i.e. twins in all pairs with both twins alive, resident in Finland and enrolled in normal public schools. Parents and teachers of the twins were also questioned in wave 1. The twins received similar questionnaires at ages 14, 17 and 22 (waves 2–4; Figure 1). The questionnaires related to the twins’ social environment (family, school, peers, neighbourhood and community), health behaviour and lifestyle factors. See Figure 1 for more details on the timing of data collection.

**Figure 1.**
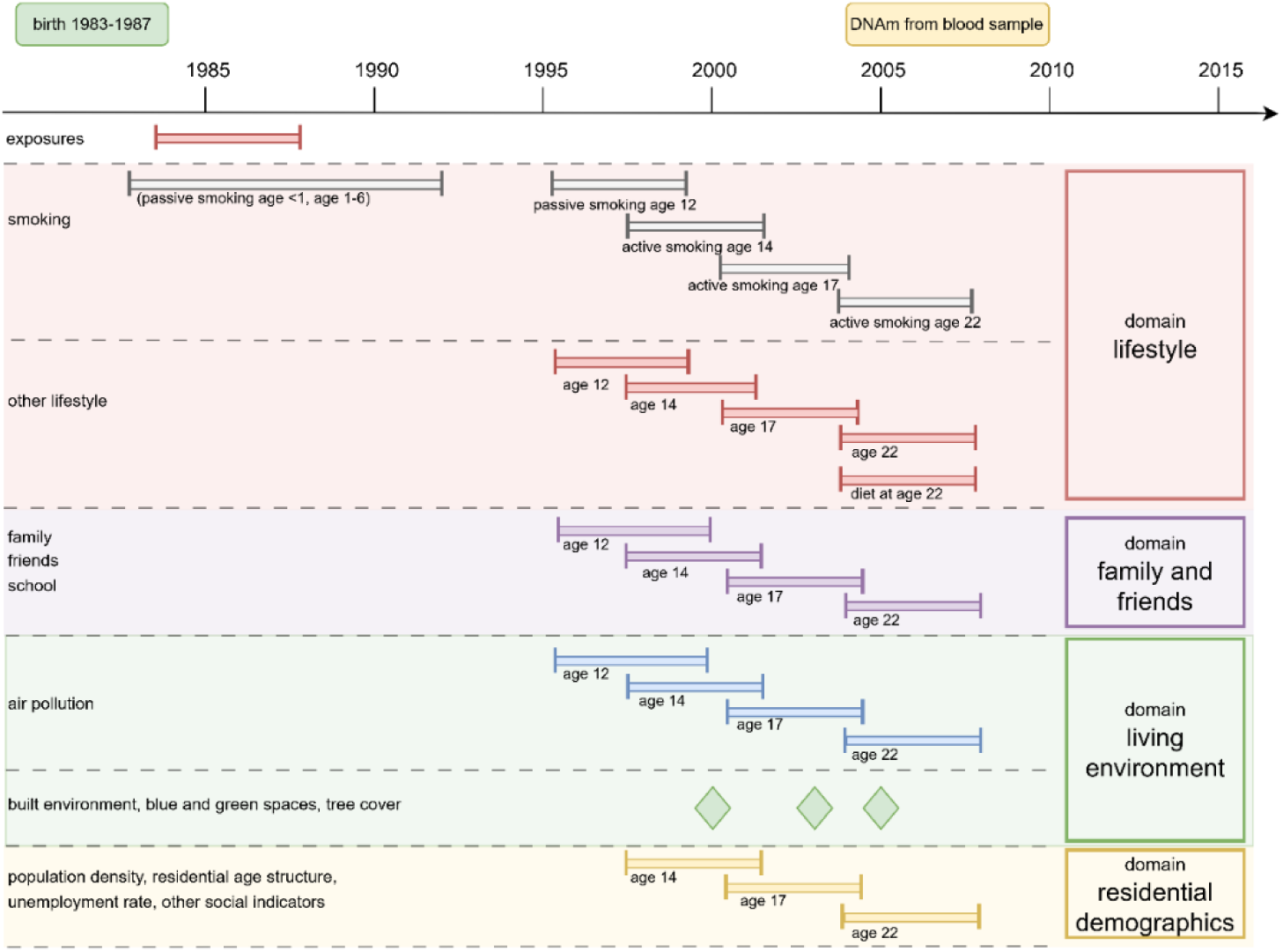
Data availability for FinnTwin 12 across the exposure domains (including all data sources).

Of the 5,184 twins who entered the study at age 12, a subsample (n=1,347) underwent intensive examinations at age 22 (mean age 21.9; wave 4), which included among other measures blood sampling and a diet survey. Our study included 848 individuals from this subsample for whom genotype, and DNAm profiles were available. After one outlier for EA (±5 SD) was removed, 847 individuals of 379 complete twin pairs and 89 single twins (i.e. the other twin was not studied) entered the final analyses.

The FinnTwin12 study was approved by the Institutional Review Board of Indiana University and the ethics committees of the Department of Public Health at the University of Helsinki (most recent HUS/2226/2021) and that of the Helsinki University Central Hospital District (HUS) (113/E3/2001 and 346/E0/05). All participants and their legal guardians provided written informed consent. The authors affirm that all procedures involved in this work adhere to the ethical standards set by the relevant national and institutional committees on human experimentation, as well as the Helsinki Declaration of 1975, revised in 2008.

### 2.2 Epigenetic Age Measurements

The primary outcome in this study is acceleration and pace of epigenetic ageing at age 22 estimated by two epigenetic clocks. We refer to both measures as EA estimates. Blood-based DNAm profiles were obtained using Illumina’s Infinium HumanMethylation450 (786 samples) or Infinium MethylationEPIC Bead Chips (93 samples) (Illumina, San Diego, CA, USA). Kankaanpää et al. (2022) describe the collection, preprocessing and normalization of DNAm data in more detail. We assessed EA utilizing two clocks selected for their superior reliability to earlier generations: the principal component-based PCGrimAge (Higgins-Chen et al., 2022; Lu et al., 2019) and the longitudinal DunedinPACE (Belsky et al., 2022). PCGrimAge utilizes 1030 CpG sites to estimate time-to-death by combining contributing factors such as chronological age, sex, smoking history and DNAm-based surrogate markers associated with seven plasma proteins (Lu et al., 2019). Age acceleration is represented by the residual arising from regressing the estimated EA (PCGrimAge estimate) onto chronological age. The DunedinPACE clock was developed on longitudinal data of one cohort in young adulthood resulting in minimized sensitivity for short-term biases and increased accuracy (Belsky et al., 2022).

### 2.3 Exposome Assessment

The exposome derived for this study consists of 556 exposures spread across ages 12 (135 exposures), 14 (166), 17 (129) and 22 (126). In all, 71 of the exposures were repeated measures through all age classes. Individual socioeconomic and lifestyle exposures derive from the FinnTwin12 questionnaires as described in Rose et al. (2019). Environmental exposures have been curated under the Equal-Life project (Julvez et al., 2021; Kamp et al., 2022) and stem from multiple sources. These were obtained by linking EUREF-FIN geocoordinates to the twins’ home addresses at specific time points (waves, Figure 1). Addresses were obtained from the Digital and Population Data Services Agency in Finland (Digital and Population Data Services Agency, 2025). To create individual urban exposomes, Statistics Finland, (2025) provided general social indicators of the living area (either at the neighbourhood postal level or for the local urban or rural community), such as crime rates, voting patterns, employment rates and income profiles. Air pollution data were acquired using previously developed air quality surfaces for Western Europe (de Hoogh et al., 2018). Exposures in this study are in the following domains: lifestyle, family and friends, living environment and residential demographics (Figure 1). They are of mixed type, i.e. continuous, binary or categorical. An exhaustive list of the exposures included can be found in the supplement (eTable 1).

### 2.4 Covariates

The choice of covariates is based on previous literature. Six covariates were determined a priori: age (whole years between birth and blood sampling), sex (male, female), zygosity (MZ, DZ), maternal age (years at time of delivery), and parental education. At wave 1, parents provided their level of primary school education (ordinal scale ranging from 1 to 6, with 6 being secondary school completion) and their highest ongoing or completed professional education (ordinal scale ranging from 1 to 5, with 5 being a university degree). Parental education is represented by a combination score, i.e. the sum of levels of school and professional education (higher score=higher level of overall education).

### 2.5 Data Preprocessing and Imputation

Participants were included in the analyses if a) complete information on outcome, covariates and full residential history was available, b) twins and their family completed at least one questionnaire at any wave and c) the missing value percentage among exposures was less than 30%. Missing data are an inherent challenge in exposome research, and in exploratory studies such as this, we deem it utmost critical to maintain multivariate relationships between exposures and avoid possible bias caused by a complete-case approach. The missingness of 22 environmental exposures was ∼50 %, because the European Urban Atlas (*European Urban Atlas*, 2025) provides reliable data only for urban and not rural areas. Detailed information on missing data and excluded exposures is provided in the supplementary material (eTable 2; eFigures 1–4). Missing general (environmental) or individual (e.g. parental exposures) data for single twins were substituted with their co-twin’s data if available, given shared residential history. Remaining missing values were imputed by performing multivariate imputation by chained equation (MICE) using the Random Forest for continuous and the Classification and Regression Trees (“cart”) method for categorical variables; *familyid* was added as a random effect to account for relatedness. Data of each wave, i.e. ages 12, 14, 17, and 22 were imputed separately to acknowledge the longitudinal structure of the dataset. We established five imputed datasets per age class with 10 iterations of chained equations each. To harmonize data across time points and ensure consistent ordering, several categorical variables were regrouped for subsequent analyses.

**Figure 2.**
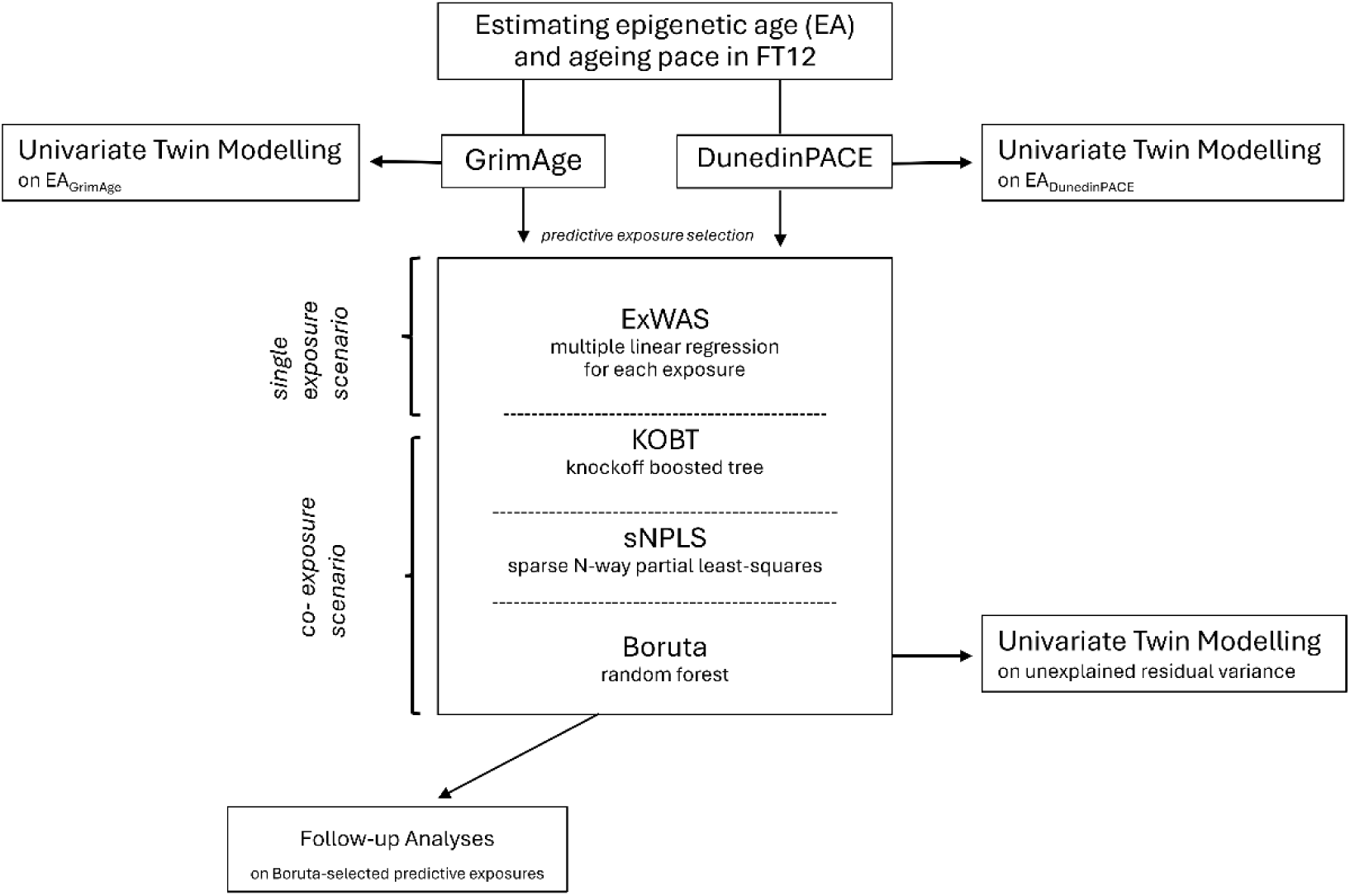
Statistical analyses performed separately on the outcomes of two epigenetic clocks (PCGrimAge and DunedinPACE).

**Figure 3.**
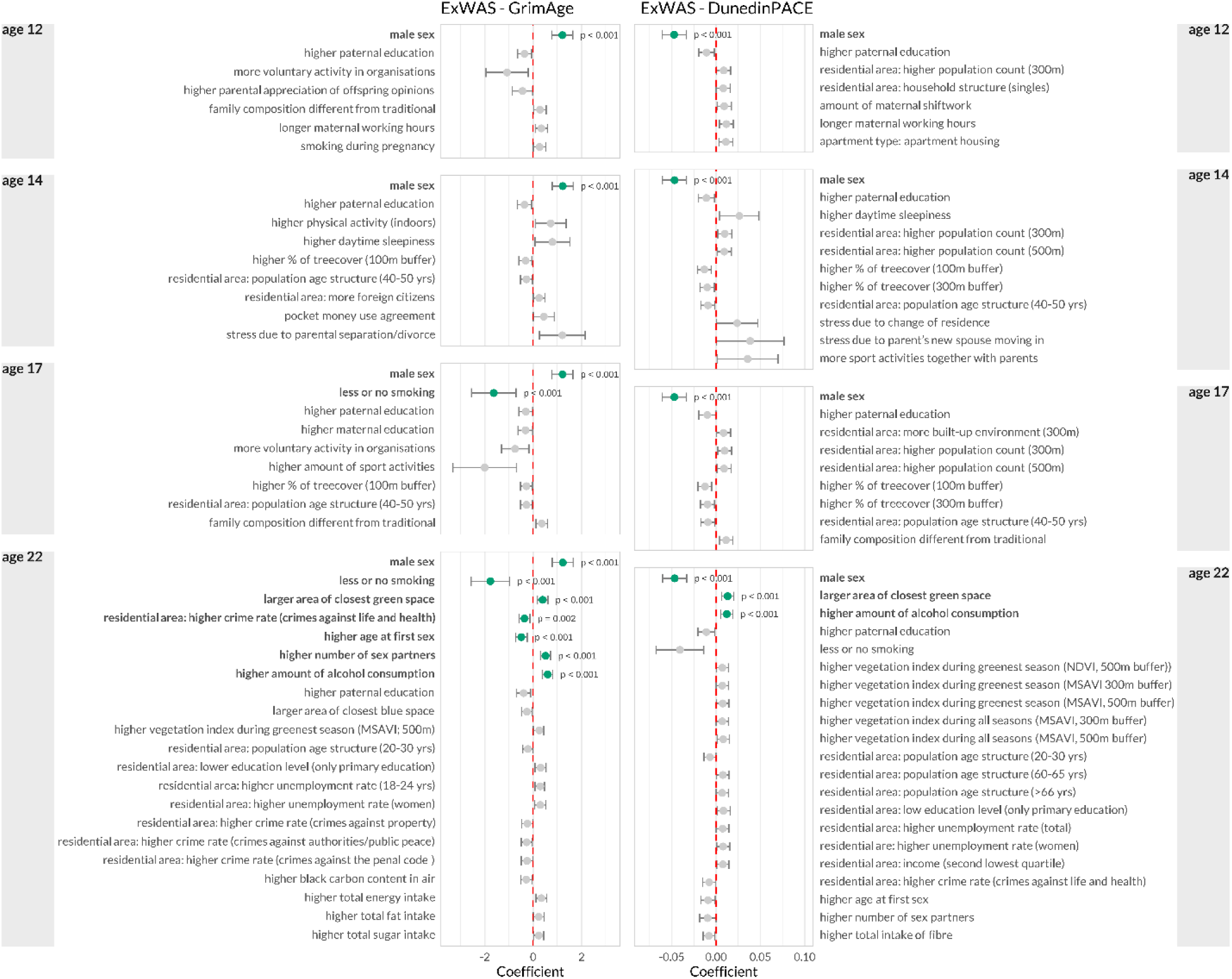
Forest plot for ExWAS results (adjusted β) for significant (FDR-adjusted <0.5, bold) and nominally significant exposures separated by age for EA_GrimAge_ and EA_DunedinPACE_; adjusted covariates were chronological age, sex, zygosity, maternal age at birth and parental education.

**Figure 4.**
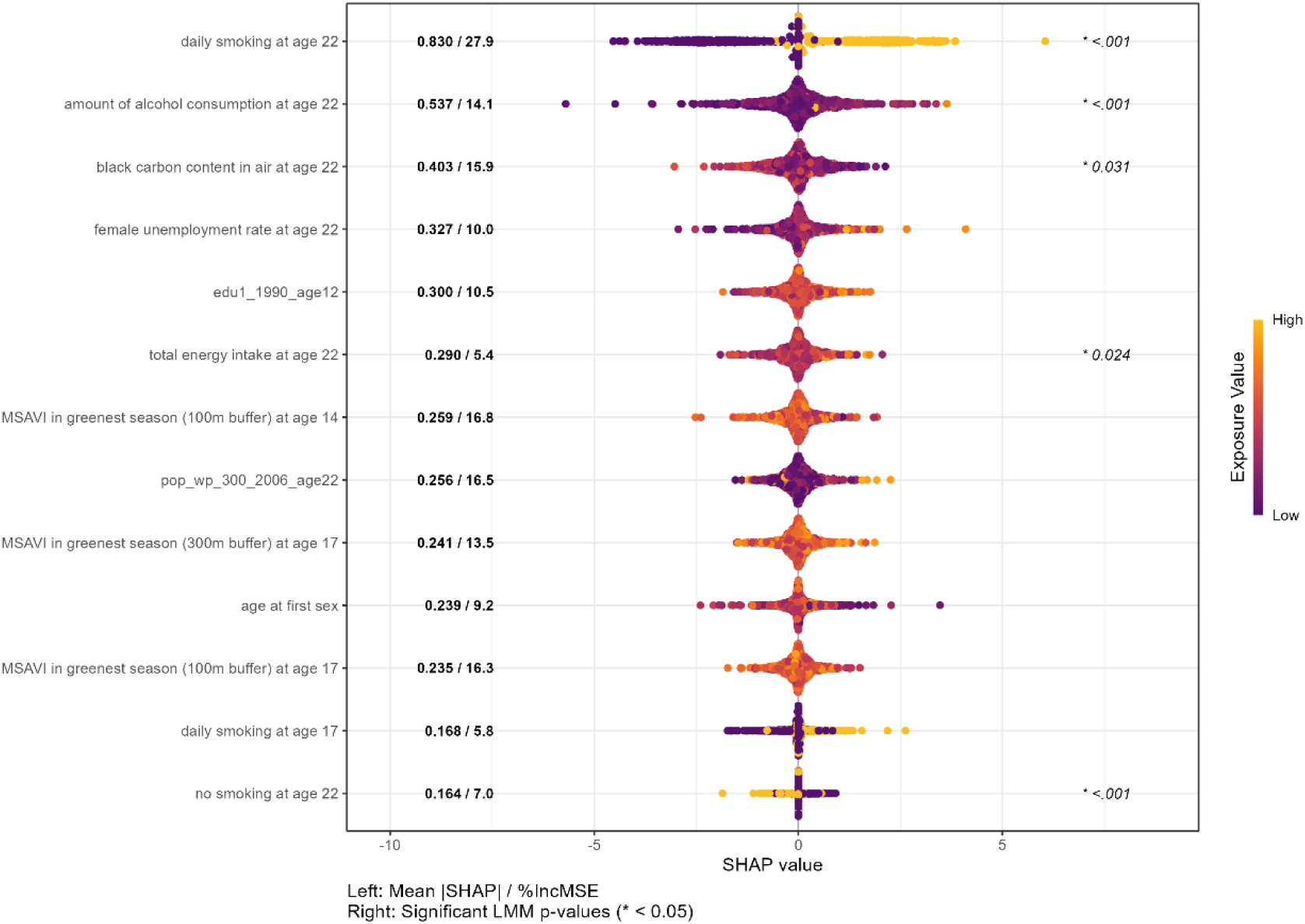
Predictive exposures of EA_GrimAge_ selected by the Boruta Stability Selection analysis (bss), ranked by SHAP value. Numbers to the left of the beeswarms are mean absolute SHAP values (mean |SHAP|) and importance scores (%IncMSE). Significant (p<0.05) results of the linear mixed-effects model (LMM) applied on this set of exposures are shown to the right of the beeswarms.

### 2.6 Statistical Analyses

#### 2.6.1 Predictive Exposure Selection

We assessed the associations between the exposome and two measures of epigenetic ageing—PCGrimAge and DunedinPACE – using several statistical approaches that either filtered for associated exposures (regression) or identified exposures important in predicting the outcome (ML models) (Figure 2). All exposure selection analyses were applied on all five subsets of MICE-imputed data and if necessary pooled afterwards using Rubin’s rules (Rubin, 1987).

##### 2.6.1.1 Single-Exposure Scenario

First, we performed two separate ExWASs on the EA estimates obtained by PCGrimAge and DunedinPACE, i.e. fitted linear mixed-effects models (LMMs) on each single exposure and adjusted the results for multiple testing by the strict Bonferroni and the less conservative Benjamini–Hochberg (FDR) corrections. Numeric exposures were standardized (mean=0, SD=1) to ensure model stability and family relatedness of twin pairs was controlled for by including a random intercept for *familyid*. All models were adjusted for the following fixed covariates: sex, zygosity, maternal and paternal education, maternal age at birth, and chronological age at assessment.

##### 2.6.1.2 Co-Exposure Scenario

The Knockoff Boosted Tree (KOBT) method performs model-free feature selection with exact FDR control (Jiang et al., 2021). In contrast to the ExWAS, KOBT can capture both linear and non-linear relationships and interactions between exposures and outcomes. The analysis was run in two modes, i.e. a) longitudinally (grouping exposures over time) and b) cross-sectionally (treating each time point independently). Repeated exposome measurements are a distinct strength of our dataset as they allow for determining relevant time windows of exposures, though increases in dimensionality and multicollinearity complicated the methodological approach. Thus, we chose to also fit a sparse N-Way Partial Least Squares (sNPLS) model to perform an alternative multivariate feature selection analysis, that explicitly models the time dimensions of the longitudinal data. This method is a multilinear regression algorithm that analyses relationships between exposures in three-way data (individuals–times–exposures) (Hervás et al., 2018). It can identify groups of exposures that co-vary with an outcome over time. A 5-fold cross-validation was performed on each of the five imputed datasets per age class to determine the optimal model parameters, specifically the number of components and the number of exposures to retain.

The final selection of predictors was based on a stability selection criterion. A predictor was considered “stable” only if it was selected (i.e. had a non-zero loading) in at least 50% of the imputed dataset analyses. sNPLS has shown good sensitivity, overall performance and interpretability but is susceptible to high FDR with increasing number of exposures and cannot handle non-linearity (Warembourg et al., 2023).

Finally, to triangulate our findings and specifically identify an ‘all-relevant’ exposure set including subtle and non-linear signals, we applied a Random Forest framework utilizing the Boruta feature selection algorithm, which ensures that correlated exposures contributing to the model’s predictive power are not arbitrarily rejected. Boruta is a wrapper method built upon the Random Forest algorithm, and, unlike standard importance ranking, it compares the true predictive power of exposures against randomized ‘shadow’ features to distinguish statistically relevant predictors from noise (Kursa & Rudnicki, 2010). The Boruta analyses (separate for the two outcomes) consisted of 100 iterations/imputation and 500 trees in each forest. A random subset of the full original and shadow predictor variables was considered at each node split.

To ensure the selected exposures were robust and not artefacts of missing data imputation, we applied a stability selection criterion, which an exposure passed if it a) ranked among the most important 50 according to their percentage increase in mean squared error (%IncMSE) and b) met this rank threshold in at least half of the imputations. Those variables were retained as Boruta-selected exposures. In a second stage, a final Random Forest regression model (500 trees) was trained using only these selected exposures. Overall model performance was evaluated using the coefficient of determination (R^2^) and at the exposure level using the %IncMSE. The latter reflects the relative importance of an exposure by estimating the loss in model accuracy if it is removed/randomized. SHAP (SHapley Additive exPlanations) values were calculated to interpret the predictive role of individual exposures. SHAP values represent the direction and strength of an exposures’ impact on prediction on the same scale as the outcome. These values inform model-level associations, and any interpretation resembling “risk” or “protective” signals arises from epidemiological interpretation rather than evidence of causality (Karimi et al., 2020; Lundberg et al., 2020).

#### 2.6.2 Follow-up Analyses

To account for the non-independence in our twin data, we fitted LMMs on the Boruta-selected predictive exposures and added *familyid* as a random intercept to control for familial clustering. This provided linear effect estimates and standard p-values to complement the ML findings.

Further, we used additional information on land use type (not included in the main models) to conduct follow-up analyses for two environmental exposures selected by the Boruta algorithm as important predictors. We investigated the association between *size of closest green space* and EA_DunedinPACE_ by fitting a linear regression model including an interaction term for *size of closest green space* and land use type. Land use was classified into eight categories: green urban areas, sport and leisure facilities, transitional woodland-shrub, mixed forests, broad-leaved forest, coniferous forest, non-irrigated arable land and land principally occupied by agriculture, with significant areas of natural vegetation. Four land use categories were dropped due to small n<10. Subsequently we examined the simple slopes (marginal trends) of green space size within each land use category (eFigure 8).

To assess collinearity between *aerial black carbon* levels and specific land use types we compared environments typical of rural and urban living using an LMM (eFigure 9). Specifically, we restricted the analysis to land use types characteristic of ‘Farming’ (non-irrigated arable land, land principally occupied by agriculture, but with significant areas of natural vegetation) and ‘Urbanity’ (sports and leisure facilities, urban green spaces), excluding all other land use types.

#### 2.6.3 Twin Modelling

To evaluate the etiological composition of variance in EA, we employed a two-stage univariate twin modelling approach based on the classical twin design. The classical twin design compares MZ twins, who are genetically identical at the sequence level, to DZ twins, who share on average 50% of their segregating genes. This comparison allows for the decomposition of variance into genetic and environmental components (Polderman et al., 2015). Genetic effects are typically divided into additive (A) and dominant (D) genetic factors. The correlation for A is set to 1.0 for twins in MZ pairs and 0.5 for twins in DZ pairs, while the correlation for D is 1.0 for MZ and 0.25 for DZ pairs. Environmental effects are divided into a common or shared environment (C), whose correlation between twins in a pair is assumed to be 1.0 regardless of zygosity, and the unique environment (E), which is uncorrelated between twins in a pair and includes unmeasured errors. Prior to the main analysis, opposite-sex DZ twins were excluded, which resolved initial violations of equal variances and means observed in EA_GrimAge_. While Shapiro–Wilk tests suggested mild deviations from normality (eTables 13 & 14), visual inspection confirmed the data were suitable for maximum likelihood estimation (eFigure 6 & 7). Assumptions of equal means and variances were met in the final sample (eTable 11 & 12). Model fit was assessed using AIC, LRT, and RMSEA, selected for its robustness to multivariate non-normality and reward for parsimony (Hu & Bentler, 1998). First, univariate twin models were used to decompose the full variance in EA estimates (PCGrimAge and DunedinPACE) into additive genetic (A), shared environmental (C), and unique environmental (E) components, which allowed us to quantify the variance explained by the exposome. In the second stage, we sought to determine the decomposition of the variance that remained unexplained by our exposure set. To isolate the variation in EA independent of the exposome, we performed non-linear residualization using another Random Forest on only the Boruta-selected predictors to calculate the residuals (the difference between observed and predicted values). We then fitted univariate twin models to the resulting unexplained residual variance. This approach allowed us to: (1) determine whether the remaining unexplained variance in EA was primarily genetic or environmental, and (2) by comparison with the baseline model, infer the extent to which the identified exposures had accounted for genetic versus environmental variance components.

### 2.7 Software

All analyses were carried out with R version 4.5.1 (R Core Team, 2025). DNAm data were processed using the R package *meffil* (Min et al., 2018) and PCGrimAge and DunedinPACE estimates were obtained through application of the following publicly available R codes: https://github.com/MorganLevineLab/PC-Clocks and https://github.com/danbelsky/DunedinPACE, respectively. MICE imputation was conducted using the R package mice (version 3.4.0; van Buuren & Groothuis-Oudshoorn, 2011). The ExWAS was performed using the lme4 package (Bates et al., 2015). KOBT is based on packages knockoff (Barber et al., 2020) and xgboost (Chen & Guestrin, 2016). Further feature selection was performed using the R packages sNPLS (Hervas, 2022), Boruta (Kursa & Rudnicki, 2010) and randomForest (Liaw & Wiener, 2002), based on the original algorithm by (Breiman, 2001). To interpret the final models, SHAP values were estimated using the fastshap package (Greenwell, 2019) and visualized using the shapviz package (Mayer, 2022).

Univariate twin modelling was conducted using structural equation modelling implemented in the OpenMx package (Neale et al., 2016).

## 3. Results

### 3.1 Study Population

Our study included 847 individuals of 379 complete twin pairs and 89 single twins from FinnTwin 12 for whom we had DNA methylation and exposome data. Of these twins, 42.3% were male. In total, 29.4% of the twins’ fathers and 43.5% of their mothers had a secondary school diploma. Of these, 16.6% of fathers and 17.8% of mothers had obtained a university degree (Table 1).

**Table 1.** Descriptive statistics of study variables by zygosity of the twin individuals (n) and of their parents (N= number of families)

|  | all twins (n=847) |  | MZ Twins (n=351) |  | DZ Twins (n=269) |  | OSDZ Twins (n=229) |  |
| --- | --- | --- | --- | --- | --- | --- | --- | --- |
|  | Mean (SD) | n | Mean (SD) | n | Mean (SD) | n | Mean (SD) | n |
| Women, n (%) |  | 491 (57.97) |  | 219 (62.39) |  | 149 (55.39) |  | 123 (53.71) |
| Men, n (%) |  | 356 (42.03) |  | 132 (37.61) |  | 120 (44.61) |  | 104 (46.29) |
| Single twins |  | 89 |  | 23 |  | 25 |  | 41 |
| chronological age, yr (4<sup>th</sup> wave) | 22.41 (0.65) | 847 | 22.43 (0.69) | 351 | 22.41 (0.71) | 269 | 22.43 (0.74) | 229 |
| PCGrimAge, yrs | 38.7 (3.23) |  | 38.8 (3.13) |  | 38.7 (3.15) |  | 38.7 (3.40) |  |
| PCGrimAge Acceleration | 0.00 | 847 | 0.06 (3.11) | 351 | -0.07 (3.13) | 269 | -0.02 (3.42) | 229 |
| DunedinPACE | 0.88 (0.10) | 847 | 0.88 (0.10) | 351 | 0.89 (0.1) | 269 | 0.89 (0.11) | 229 |
| Parental data |  |  |  |  |  |  |  |  |
| Maternal age at birth | 29.83 (4.6) | 468 | 28.93 (4.8) | 187 | 30.34 (4.6) | 147 | 31.0 (4.4) | 135 |
| Paternal age at birth | 32.06 (5.6) | 468 | 31.14 (5.58) | 187 | 32.06 (5.38) | 147 | 32.93 (5.19) | 135 |
| Maternal education level<sup>a</sup>, range 1–9 | 6.1 (2.2) | 468 | 6.0 (2.2) | 187 | 6.1 (2.3) | 147 | 6.1 (2.3) | 135 |
| Paternal education level<sup>a</sup>, range 1–9 | 5.2 (2.5) | 468 | 5.2 (2.5) | 187 | 5.3 (2.5) | 147 | 5.1 (2.5) | 135 |

<sup>a</sup> combined score of primary, secondary and adult education + vocational training (higher score=higher level of overall education)

### 3.2 ExWAS

To agnostically screen independent, single exposures’ associations with the outcome, two ExWAS analyses were performed on EA_GrimAge_ and EA_DunedinPACE_, respectively. A total of 847 individuals and 556 exposures were included. None of the exposures for either outcome withstood the strict Bonferroni correction for multiple testing and only nine exposures altogether (seven for PCGrimAge, two for DunedinPACE) remained significant after FDR adjustment (eTables 3 & 4). The covariates *sex* and *paternal education level* were significant and nominally significant, respectively, throughout all ages and both outcomes. The ExWAS analyses at age 12 and 14 revealed no exposures significantly associated with EA at age 22. Smoking was identified as the strongest positive predictor of biological ageing at ages 17 (EA_GrimAge_) and 22 (EA_GrimAge_ and EADunedinPACE), followed by the amount of alcohol consumption at age 22 (EADunedinPACE). Higher EAGrimAge was associated with younger age at sexual debut and with a higher number of sex partners. The only exposure from the ‘living environment’ domain leading to significantly elevated levels of EADunedinPACE was area of closest green space at age 22. The larger this green space was, the higher the pace of biological ageing is. However, many exposures related to the living environment, such as green space, tree cover and the vegetation index were identified by both clocks as nominally significant. Altogether, 26 (PCGrimAge) and 38 (DunedinPACE) nominally significant exposures were found (Figure 3). Some of these, such as amount of smoking, age at sexual debut and number of sex partners (all at age 22, DunedinPACE) replicated the significant associations observed for EAGrimAge; others represented new exposures from the domains of lifestyle, family and friends, living environment and residential demographics.

### 3.3 KOBT and sNPLS

To model the interplay between multiple exposures and/or repeated measures (co-exposure scenario), we applied two complementary methods to identify exposures associated with epigenetic ageing in a non-linear fashion (KOBT) and in a longitudinal structure (sNPLS). KOBT applies rigorous FDR control, and it did not identify a single significant predictor among the exposures for neither mode (longitudinal and cross-sectional) nor any outcome. The analysis provided a set of predictor exposures that are close to the significance threshold based on their importance scores (eTable 5; eFigure 5). Exploratory analysis in this set of exposures revealed a pattern of selected exposures similar to that in the ExWAS analysis. Consistently among the most likely influential predictors for both ageing outcomes and in both longitudinal and cross-sectional models were *amount of smoking, amount of alcohol consumption, low-income level in residential area and vegetation index (MSAVI).* The three covariates *sex*, *paternal education* and *maternal age at birth* were also chosen consistently as important predictors of the outcome.

**Figure 5.**
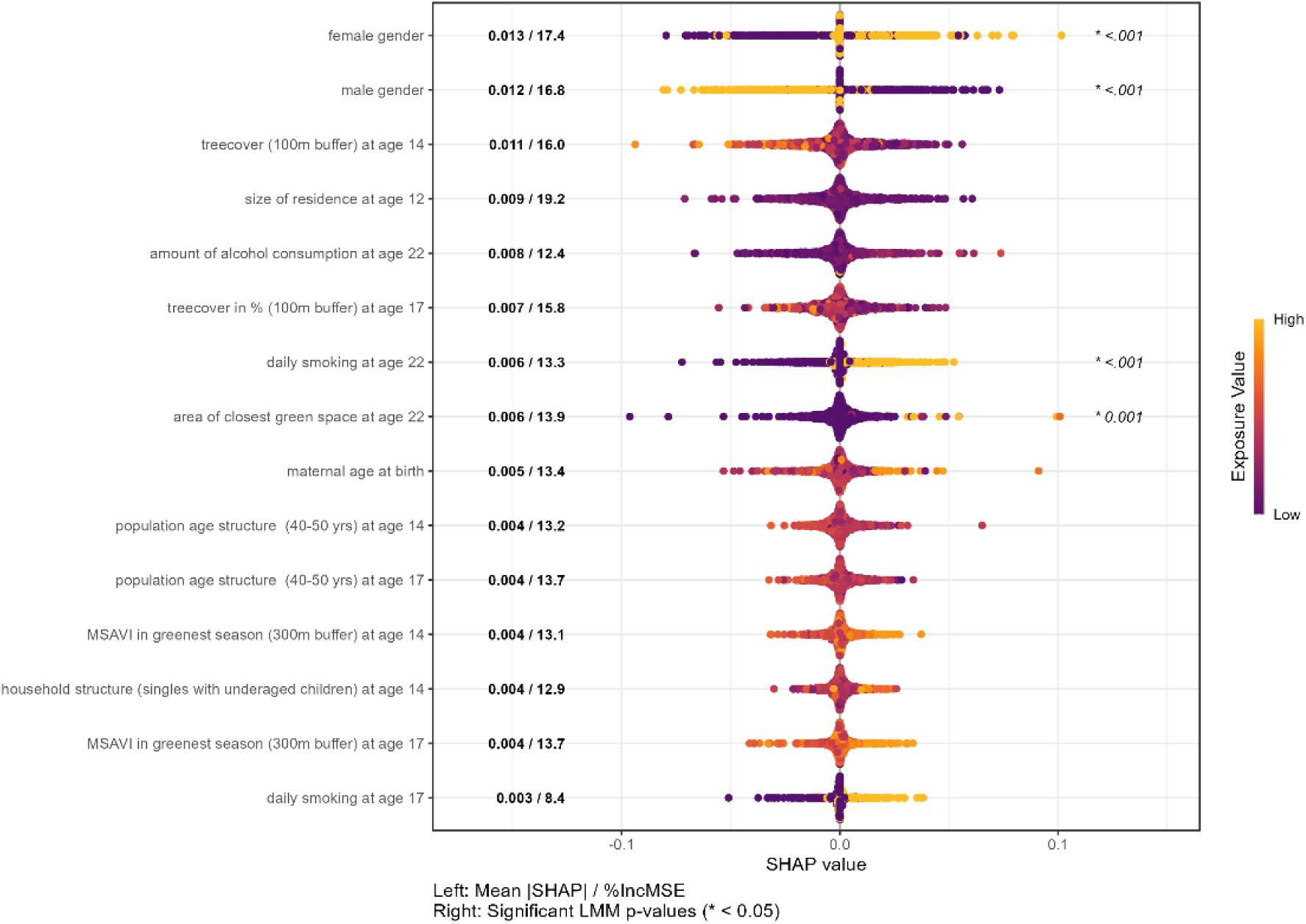
Predictive exposures of EA_DunedinPACE_ selected by the Boruta Stability Selection analysis (bss), ranked by SHAP value. Numbers to the left of the beeswarms are mean absolute SHAP values (mean |SHAP|) and importance scores (%IncMSE). Significant (p<0.05) results of the linear mixed-effects model (LMM) applied on this set of exposures are shown to the right of the beeswarms.

**Figure 6.**
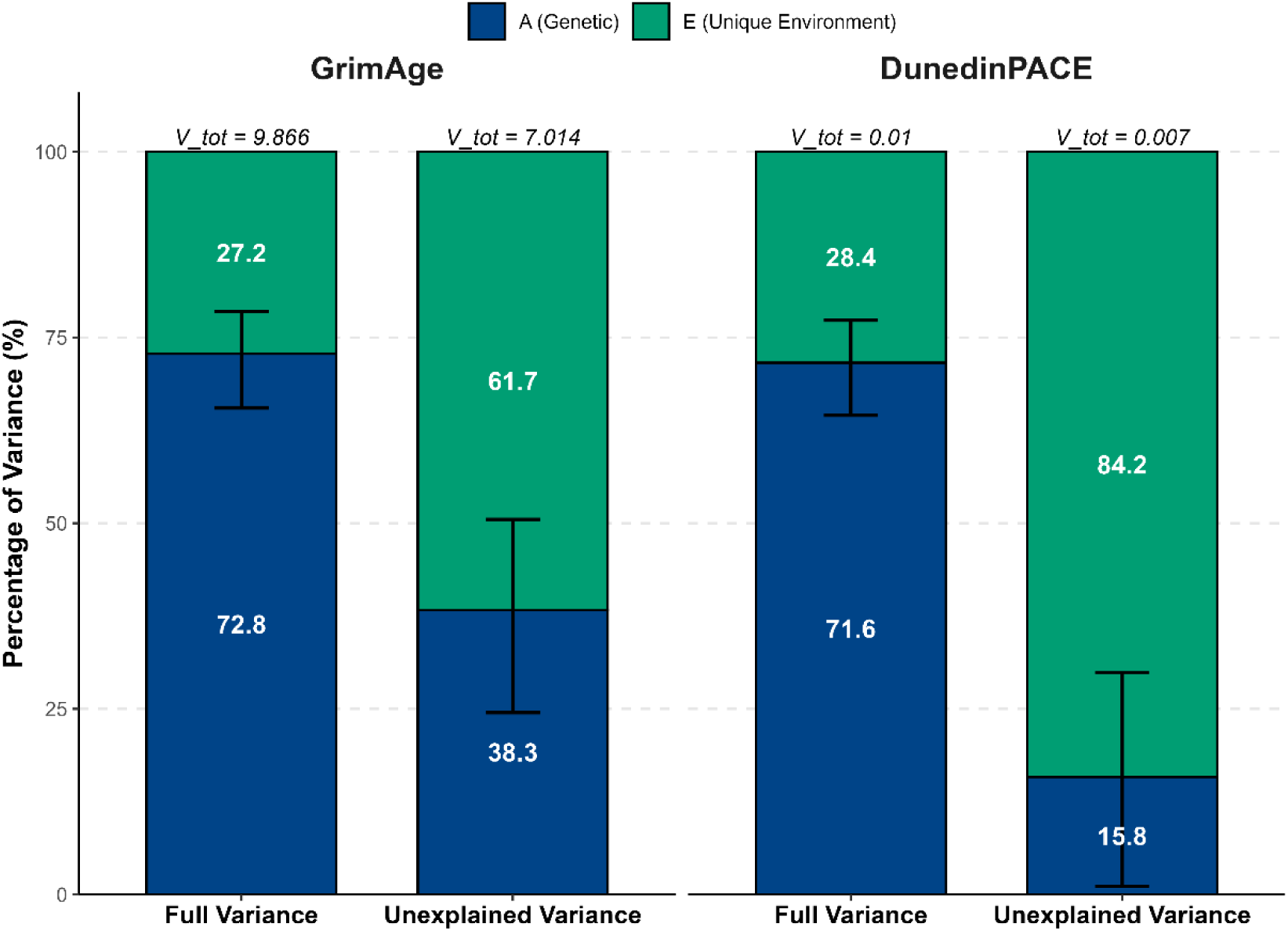
Univariate twin model (Cholesky decomposition): Standardized estimates of variance decomposition of EA_GrimAge_. Raw = total variance in EA_GrimAge_ and EA_DunedinPACE,_ respectively. Total variance (V_tot) is annotated above the bars. Unexplained variance represents variance in EA_GrimAge_ and EA_DunedinPACE,_ after adjusting for the full current exposure set (explained variance EA_GrimAge_ =25.7%, EA_DunedinPACE_=30.8%). The model was adjusted for the covariates chronological age, sex, maternal age at birth, maternal and paternal education levels.

sNPLS was conducted to find stable longitudinal predictors of EA among the exposures, but it failed to identify any passing the stability selection criterion (eTable 6).

### 3.4 Boruta Stability Selection and Linear Mixed-Effects Model

Two data-driven Random Forest analyses (Boruta) were applied to distinguish relevant predictors from noise, identifying exposures consistently important for predicting EA. The algorithm selected 13 exposures predictive of EA_GrimAge_ (explained variance R^2^=25.7, Figure 4). For EA_DunedinPACE_ (R^2^=30.8), 39 predictive exposures were identified. Figure 5 highlights the top 15 most influential predictors among them. The remaining 24 exposures contributed less substantially to prediction accuracy (eTable 7 for the full list of selected exposures). To validate the exposure selection, we compared the predictive power of the final model (including only the selected predictors) against a baseline model (including all 500+ exposures). The final model explained more variance than the baseline model for both outcomes, confirming that exposure selection successfully reduced noise (eTable 8). SHAP values quantify each exposure’s contribution to EA prediction expressed in the scale of the outcome, whereas the importance score (%IncMSE) reflects the impact of individual exposures on model performance. The Boruta analysis treated twins as independent individuals to capture maximum predictive signals (including non-linearities), while subsequent LMMs tested strictly for linear effects while accounting for the twin structure (eTables 9 & 10). Therefore, exposures with high SHAP values but lacking LMM significance likely indicate strong non-linear associations with EA, whereas LMM-significant exposures represent robust linear predictors even after controlling for family relatedness (Figures 4 & 5).

The Boruta-selected sets of predictive exposures show substantial overlap with results of the ExWAS and partially that of KOBT. Among the predictors ranking high or medium in SHAP appear those that also reached statistical significance in the ExWAS and LMM (*amount of smoking* at age 22 (mean |SHAP| = 0.841; %IncMSE = 30.7; ExWAS 1.770 (2.570, 0.971), p_adj_.=0.005), *amount of alcohol consumption* at age 22 (mean |SHAP| = 0.563; %IncMSE = 13.0; ExWAS 0.601 (0.391, 0.810), p_adj_.<0.001), and *area of closest green space* at age 22 (mean |SHAP| = 0.005; %IncMSE = 15.1; ExWAS 0.013 (0.006, 0.019), p_adj_.= 0.01)). Among the assumed non-linear effects *size of residence* at age 12 stands out in that this exposure scores high in SHAP (0.009) and is the most influential for model performance (%IncMSE = 19.7) yet was not identified by the ExWAS. In contrast, the assumed non-linear negative association of a higher *age at sexual debut* with EA_GrimAge_ (ExWAS −0.489 (−0.729, −0.247), p_adj_. =0.003) only played a minor role in predictive accuracy (mean |SHAP| = 0.220) and model performance (%IncMSE = 8.1). A higher *aerial black carbon* at age 22 ranked high in SHAP (0.375) and %IncMSE (11.9) and reached nominal significance in the ExWAS (−0.283 (−0.517; −0.048), p_unadj_. =0.018), while it was – counterintuitively – predictive of decreased EA scores.

Both epigenetic clocks additionally identified predictive demographic exposures and environmental exposures, which notably showed high importance scores throughout (PCGrimAge: range: 11.2–18.5; %IncMSE_max_: 30.7, %IncMSE_min_: 5.3) (DunedinPACE: range: 12.5–16.4; %IncMSE_max_: 19.7, %IncMSE_min_: 10), independent of their rank in predictiveness. This suggests, that while these exposures may not strongly influence EA, they are important for accurately predicting it.

Our analytical funnel identified both linear and non-linear influences linking lifestyle, environmental, and demographic factors to epigenetic ageing. Tables 2 and 3 summarize exposures that were consistently highlighted across the methods, providing a triangulated view of predictive factors.

**Table 2.** Predictive exposures of EA_GrimAge_ selected by Boruta Stability Selection analysis and at least one other method. Nominally significant exposures are presented in parentheses. Effect directions were determined by ExWAS.

| Exposure | ExWAS | KOBT | sNPLS | BORUTA | effect direction | predictive impact on EA |
| --- | --- | --- | --- | --- | --- | --- |
| daily smoking at age 22 | ü | ü |  | ü | positive | increases |
| higher amount of alcohol consumption at age 22 | ü | ü |  | ü | positive | increases |
| higher aerial black carbon at age 22 | (ü) |  |  | ü | negative | decreases |
| residential area: higher unemployment rate (18-24 yrs) at age 22 | (ü) | ü |  | ü | positive | increases |
| higher population density (500m buffer) at age 14 | (ü) | ü |  | ü | positive | increases |
| total energy intake at age 22 | (ü) |  |  | ü | positive | increases |
| higher age at sexual debut | ü |  |  | ü | negative | decreases |
| no smoking at age 22 | ü | ü |  | ü | negative | decreases |
| daily smoking at age 17 | ü | ü |  | ü | positive | increases |

**Table 3.** Predictive exposures of EA_DunedinPACE_ selected by Boruta Stability Selection analysis and at least one other method. Nominally significant exposures are presented in parentheses. Effect directions were determined by ExWAS.

| Exposure | ExWAS | KOBT | sNPLS | BORUTA | effect | predictive impact on EA |
| --- | --- | --- | --- | --- | --- | --- |
| higher tree cover in % (100m buffer) at age 14 | (ü) | ü |  | ü | negative | decreases |
| higher tree cover in % (100m buffer) at age 17 | (ü) | ü |  | ü | negative | decreases |
| daily smoking at age 22 | (ü) | ü |  | ü | positive | increases |
| large size of closest green space at age 22 | ü |  |  | ü | positive | increases |
| higher vegetation index during greenest season (MSAVI, 300m buffer) at age 14 |  | ü |  | ü | negative | decreases |
| residential area: population age structure (more 40-50yr olds) at age 17 | (ü) | ü |  | ü | negative | decreases |
| residential area: population age structure (more 40-50yr olds) at age 14 | (ü) |  |  | ü | negative | decreases |
| higher vegetation index during greenest season (MSAVI, 300m buffer) at age 17 |  | ü |  | ü | negative | decreases |
| daily smoking at age 17 |  | ü |  | ü | positive | increases |

#### 3.4.1 Follow-up Analyses

A larger *size of closest green space* at age 22 was identified as a primary factor positively associated with increased acceleration in biological ageing (EA_DunedinPACE_; β= 0.012, 95% CI [0.005, 0.018], p < 0.001). Although our post-hoc test on overall interaction between this exposure and type of land was not statistically significant (F {15,801} = 1.24, p= 0.24), an exploratory analysis of simple slopes suggested a significant positive association solely with *non-irrigated arable land* (β= 0.011, 95% CI [0.000, 0.023], p = 0.045; eTable 17; eFigure 8). The linear mixed model on correlations between *aerial black carbon* levels and specific land use type showed that black carbon levels were significantly higher in the urban environment (sports and leisure facilities, urban green spaces) compared to the ‘farming’ environment (β= 0.26, 95 % CI [0.19, 0.34], t=6.72, p < 0.01; eTable 18; eFigure 9).

### 3.5 Twin Modelling

To assess the nature of the variance captured by our exposure set and the unexplained residual variance, we fitted two separate univariate twin models: one on the unadjusted (full) EA (eTable 15) and another on the variance left unexplained by our exposure set (eTable 16). Comparing these models allowed us to quantify how much of the explained variance was attributable to shared genetic factors (A, indicating gene–environment correlation) versus unique environmental effects (E).

Our univariate twin models (Figure 6) show, that for EA_GrimAge_, genetic factors (A) explained the majority of variance in the full measure (73%), with unique environmental factors (E) accounting for 27% (V_tot = 9.87). After adjusting for the full set of exposures available and fitting another univariate twin model to the variance left unexplained by our exposure set, we detected a clear shift in composition. We found that this remaining variance (V_tot = 7.01) is predominantly driven by unique environmental factors (62%), with a substantially reduced genetic component (38%) compared to the unadjusted model. For EA_DunedinPACE_, the full variance showed a similar pattern, with genetic factors explaining 72% and unique environment 28% (V_tot = 0.01). After adjustment, the genetic component in unexplained variance dropped to 16%, and the unique environment component rose to 84% (V_tot = 0.007). The difference in explained variance between the Boruta and the twin models (EA_GrimAge_ 25.7% vs. 27%; EA_DunedinPACE_ 30.8% vs. 38%) reflects different approaches of predicting versus decomposing the variance in EA. The twin model was performed on the full set of exposures (including noise) adjusted for the twin structure in our data, but it cannot capture non-linear effects as the Boruta can. Also, measurement error cannot be accounted for in either of the models.

Despite the small difference, the close alignment in magnitude of explained variance (25%–30% vs. 27%–38%) between these conceptually distinct models (prediction vs. decomposition) indicates that the Boruta-selected exposures capture a substantial and consistent signal related to epigenetic ageing, reinforcing their robustness as predictors.

## 4. Discussion

Our study presents a selection of exposures predictive of epigenetic ageing in young adulthood and a genetically informed evaluation of the exposome set we applied. We integrated multiple statistical frameworks to achieve robustness of predictors to methodological differences. Furthermore, we leveraged univariate twin models to gain information on the coverage and composition of EA variance in our exposome data as well as the unexplained, “missing” part. In the FT12 cohort, we identified nine consensus-derived exposures predictive of EA_GrimAge_ and nine partially overlapping predictors of pace of ageing (EA_DunedinPACE_). The identified predictors explained approximately 30% of EA variance, with a substantial proportion of this variance attributable to genetic factors. No single exposure domain was dominant, as predictors ranged from lifestyle (smoking, alcohol, energy intake and sexual debut) over demographics (unemployment rate, population density and age structure) to general environment (tree cover, vegetation index and air pollution). Our results further highlight that non-linear dynamics and co-exposure interactions are crucial in EA prediction, as they reflect biological complexity that linear methods fail to model. This is particularly relevant given our finding that most of the unexplained residual variance in EA is attributable to unique environmental (non-shared) exposures. The two epigenetic clocks show similarity in that both predictor sets include factors from lifestyle, environment and demographics. It is notable however, that they specifically align only in two exposures, i.e. identifying *daily smoking at ages 17 and 22* as positive predictors of EA. This limited overlap (2/18 predictive exposures) is not indicative of inconsistency but rather highlights the differences between the measures, e.g. the accumulated state of damage (PCGrimAge) versus the current rate of physiological decline (DunedinPACE). The convergence of both clocks on the negative impact of daily smoking underscores its potency as a systemic toxicant, although the higher importance in the PCGrimAge and lower score in the DunedinPACE model reflect what those clocks were trained on. PCGrimAge estimates are based on mortality risk, specific plasma proteins and smoking pack years, thus favouring lifestyle exposures, that exert effects on inflammatory and mortality biomarkers (Lu et al., 2019). In contrast, DunedinPACE was developed to capture longitudinal or chronic stressors/buffers that have a subtle but steady impact on EA, such as general environmental exposures (Belsky et al., 2022). There are also substantial differences in the life circumstances of 17-year-olds and young adults who had a mean age of 22 in the present study.

### 4.1 Predictive Exposures

Within the ‘lifestyle’ domain, our results on smoking and alcohol consumption as positive predictors of higher EA are not surprising and support earlier studies (Kankaanpää et al., 2025; Kresovich et al., 2022). The association between a *higher age at sexual debut* and lower EA also supports former findings (Schlomer, 2024; Zhang et al., 2023). BMI played a mediating role in the latter study and was also included in our set of exposures (at age 22), but it was not among the identified predictors of EA. Considering exposure interaction, a lower *age at sexual debut* could be indicative of early-maturing youth, who are more susceptible to e.g. alcohol consumption and smoking due to pubertal stress, underdeveloped self-regulation and sensation-seeking (Dick et al., 2000; Goering et al., 2024; Kong et al., 2013). The identification of *total energy intake at age 22* as a positive predictor of EA in absence of any BMI impact in our study is interesting, as it is initially unexpected regarding earlier positive associations between BMI and EA (Kresovich et al., 2022; Ravi et al., 2025). However, these previous observations are possibly driven by insulin resistance rather than a direct effect (Lundgren et al., 2022). We further assume that *total energy intake* interacts with alcohol consumption and more subtly with dietary behaviour and socio-demographic exposures (e.g. education or income level).

It is logically sound that macro-level factors within the ‘residential demographics’ domain – such as youth unemployment rates, population density and age structure – emerged as EA predictors, given that individual health is embedded within a broader socioeconomic context. It is remarkable, though, considering that Finland’s social equality is considered rather high and neighbourhood differences in SES are not as distinct as elsewhere (Viinikka et al., 2023). The identified positive association of higher neighbourhood *unemployment rate (18–24 yr olds)* with EA_GrimAge_ confirms the sensitivity of our approach to age-specific contexts as it represents the twins’ specific peer group at age 22. A higher number of 40–50-year-olds in the residential area at ages 14 and 17 associates with lower EA_DunedinPACE_ in our study. Since that age group likely represents the parental generation of the twins, we assume this could reflect a more stable, family-oriented neighbourhood environment that provides a greater sense of safety for adolescents. Demographically, this age group is associated with established economic resources and higher residential stability (Sampson & Raudenbush, 1999). Furthermore, the presence of established families implies a higher density of local, age-matched peers proven to be crucial for adolescents’ health and wellbeing (Meyers et al., 2013; Viner et al., 2012).

Regarding the ‘living environment’ domain, our results demonstrate the importance of environmental exposures as chronic stressors or buffers influencing EA_DunedinPACE_. These exposures might exert a small yet constant effect size over time – the type of cumulative impact that DunedinPACE is designed to capture. Although Nordic environments are generally characterized by abundant greenness, *tree cover* and *vegetation index* still contributed distinctly to explaining variance in EA. Their protective qualities have been shown in previous exposome studies (Barboza et al., 2021; Markevych et al., 2017; Münzel et al., 2023). Counterintuitively, our results show a positive association of *size of closest green space* with EA_DunedinPACE_, contradicting the standard ‘green space’ hypothesis and previously found protective effects against e.g. telomere shortening, another biomarker of ageing (De Ruyter et al., 2022). This discrepancy may be attributed to the nature and land use of those spaces. In Finland, large green areas often signify rural remoteness rather than recreational urban parks (Wang et al., 2025) and we could demonstrate that increased EA_DunedinPACE_ due to larger *size of closest green space* interacts significantly with the land use type *non-irrigated arable land*, indicative of rural, agricultural living. Forest lands were not associated with increased EA. Previous studies indicate that while urban green space promotes physical activity, vast rural green spaces can be associated with car dependency, longer distances to services and sport hobbies, and lower walkability of living environments (Kleinert & Horton, 2016; Molina-García et al., 2020; Sallis et al., 2016). Evidence in children are inconsistent, however (Aznar et al., 2024; McCrorie et al., 2021). Our findings align with those of Cardinali et al. (2024), who reported negative indirect health effects of surrounding greenness within certain buffers and green space use. They argue that green spaces must be accessible to benefit health, but in rural areas these are often private or agricultural land. In contrast to previous research, we observed an inverse association where higher *aerial black carbon* associated with lower EA_DunedinPACE_ scores (Baccarelli et al., 2009; Hoek et al., 2002). This finding likely reflects the ‘urban advantage’ effect, rather than a true benefit, as our post-hoc test also showed that high black carbon levels were associated with land use categories typical of urban environments (eTable 18; eFigure 9). In Nordic countries, unlike many other regions, higher air pollution exposure is positively correlated with higher SES, education and income (Strandell et al., 2024). Likewise, lower SES and higher unemployment rates are linked to low population density indicative of rural living (Bremberg, 2020; Weeks et al., 2023). The neighbourhood socioeconomic disadvantage in rural areas is associated with higher alcohol consumption, poorer diet, less physical activity; and higher rates and earlier onset of smoking (Dick et al., 2009; Egan et al., 2024; Kivimäki et al., 2022; Lagström et al., 2022). Earlier twin models showed a higher impact of environmental influences on adolescents’ drinking behaviour in rural areas compared to urban settings where the genetic background plays a more substantial role especially in girls (Dick et al., 2009; Rose et al., 2001). Consequently, the apparent negative association of *aerial black carbon* with EA_GrimAge_ may be a statistical artefact acting as a proxy for higher living standards in urban centres and the negative impact of large green spaces vice versa for neighbourhood disadvantages in rural settings.

The notable lack of individual lifestyle exposures predictive of EA – beyond smoking, alcohol use and sexual debut – is surprising, given that these exposures reflect the individual environment where twins might differ. However, adolescents rarely exhibit a consistent lifestyle or experience a steady social environment. In general, adolescence and young adulthood imply turmoil and frequent changes in social exposures (school changes, moving out, friendship turnover, etc.), possibly adding random effects to our data.

### 4.2 Twin Models

Beyond identifying EA predictors, we sought to evaluate the exposure set we applied by utilizing the twin structure in our data. Applying a classic twin design offered insight into the nature of our exposure set and likewise the “missing or unexplained” exposome. The epigenetic ageing measures exhibited similar trends in these analyses, which is why we discuss them in general. In the baseline model, EA was largely genetic (A∼70%), with the unique environment (E) explaining only ∼30%. However, after adjusting for our exposure set (explaining ∼ 27% of total variance), the unexplained residual variance showed the opposite pattern: genetic contribution diminished to ∼38%, while unique environmental exposure’s share rose to ∼62%. This implies that our exposome set successfully modelled a substantial proportion of the genetic component in epigenetic ageing. We infer that our identified predictors are driven by a mix of A and E, consistent with previous studies on e.g. education, unemployment and neighbourhood composition (Fiorito et al., 2017; Freni-Sterrantino et al., 2022). This aligns with the concept of passive gene–environment correlation, where parents shape the living environment of their children based on their own genetic predispositions (Kandler et al., 2024). During early adulthood, the impact of individual environmental factors increases, as twins become more autonomous or cohabitation with their co-twin ends. Many of the participating twins were still living in the same household at age 22 or had just recently moved apart from each other (Wang et al., 2022). They were, however, likely at the verge of this transition into a more self-directed life; thus, a follow-up on EA is needed to observe this effect more clearly.

The unexplained variance is predominantly determined by unique, non-shared environmental exposures. This composition suggests that the unexplained drivers of EA likely include unmeasured environmental variables with a low genetic component, random individual exposures (e.g., adverse life events), stochastic biological noise including epigenetic drift, and measurement error. Further, we assume, that other omics layers and/or environmental toxicants, which our study did not include, may represent critical components of our estimated missing exposome. Chemical pollution alone is estimated to account for approximately 16% of global deaths, which underscores the potential significance of these exposures in shaping EA (Landrigan et al., 2018).

### 4.3 Strengths and Limitations

Only a few studies have investigated associations between a large range of longitudinal, differing domain exposures and epigenetic ageing clock estimates using agnostic statistical methods (e.g. de Prado-Bert et al., 2021; Khodasevich et al., 2025), and to our knowledge ours is the only one to include twin modelling.

The major strength of our data-driven study is the unique combination of an extensive twin dataset and an analytical strategy that proceeds from initial screening of single exposures to non-linear patterns predictive of EA. Our approach addresses challenges related to studying a) biological ageing in adolescents and young adults and b) the human exposome in its complexity. With our twin model we address a serious shortcoming in current exposome research regarding the ambiguity in scope. Further, we are not aware of any study that empirically assessed the etiological composition of the specific exposure subset they investigated.

We publish all exposure–EA associations to avoid selective reporting (Reid et al., 2015). Each method in our study has its benefits and disadvantages, but combined they provide a robust picture. While ExWASs provide an unbiased screen, they are inherently broad and require rigorous downstream validation to separate signal from noise. They are highly sensitive to false discoveries and correlated exposures, and our sample size was likely insufficient for strict Bonferroni correction (Chung et al., 2019). Multicollinearity likely contributed to the lack of significant findings in our KOBT analysis, as signals were dispersed across correlated exposures. We view this not as a flaw but as evidence of the exposome’s complexity. While previous studies have often addressed multicollinearity by manually pooling or dropping all but one of the correlated exposures (e.g. buffer zones), we consider this practice inconsistent with the exposome concept. Instead, we applied the data-driven Boruta method, which accounts for multicollinearity while robustly distinguishing signal from noise. Interactions between exposures – central to the exposome idea – remain underexplored despite an increasing trend towards multi-exposure studies (Maitre et al., 2022). Overfitting and leakage are potential issues in our study, since all analyses used the same unsplit dataset. However, feature selection methods were mutually independent, and their observational units (individuals) differed from those of twin models (twin pairs). We identified predictive exposures within the FT12 cohort, but replication in an independent cohort (of twins or of non-twin singletons) is needed to confirm their validity and generalizability. Nonetheless, our results should be considered a robust source of hypothesis generation. In addition, the findings should be interpreted in light of the challenges of measuring biological ageing in young adults, where age-related biological differences might not yet be distinct. Yet, despite a slow rate of DNAm change (∼2%–5% over many decades; (Seale et al., 2024), the accuracy of the clocks is high (Teschendorff & Horvath, 2025) and their estimates bare importance considering developmental plasticity. It is still unclear what molecular mechanisms exactly drive them, and their algorithms differ in terms of the parameters and tissue used for training them. We chose to conduct our study with different clocks sensitive to cumulative (PCGrimAge) and short-term (DunedinPACE) environmental exposure effects, both of which are highly validated in younger cohorts (Kuznetsov et al., 2025). This way we ensured coverage of past as well as current environmental effects. The overlapping results between the clocks strengthen our findings. Our twin models may have been subject to inherent estimation biases due to the fact, that many twins were still sharing an address at age 22 or had only recently moved apart, which in the primary model on exposome-adjusted variance in EA might have slightly inflated the additive genetic component (*A*), as in *AE* models it absorbs all shared environmental influences that lack a corresponding *C* term. Furthermore, the *E* component in our model on residual, unexplained variance likely represents an upper-bound estimate of unique environmental impact, as it inherently incorporates both stochastic biological noise and any remaining measurement error. A follow-up study in the FT12 cohort, after the twins have lived apart from each other for a longer time would yield more accurate estimates. To validate and extend our approach and findings, future research should integrate multi-omics and expanded exposure datasets, leverage causal inference methods to test mechanisms, and ensure replication across diverse independent cohorts.

## 5. Conclusion

We identified robust predictors explaining the variance in epigenetic ageing in young adulthood, ranging from established lifestyle risks like smoking and alcohol use to negatively associated factors such as exposure to greenness and age-appropriate residential demographics. The evidence for non-linearity and interactions is particularly critical, as it suggests that simple effects models would fail to capture most of the unexplained variance of mainly unique environmental exposures. Our twin modelling of the unexplained variance in EA (“missing exposome”) demonstrates that our exposome set effectively captured the shared genetic and environmental influences, leaving a remainder dominated by unique, non-shared environmental factors that drive individual differences within families. Early life exposures program later life health, and our study identified modifiable systemic factors. These offer a societal lever to precision interventions targeting health equity at the population level beyond what can be achieved in targeting individual behaviour alone.

## CRediT authorship contribution statement

Annika Opperbeck: Writing – original draft, Methodology, Formal analysis, Conceptualization, Data Curation, Software, Visualization, Funding acquisition. Zhiyang Wang: Writing – review & editing, Validation, Methodology, Conceptualization, Data curation. Ilkka Rautiainen: Writing – review & editing, Methodology, Validation. Aino Heikkinen: Writing – review & editing, Data Curation. Jaakko Kaprio: Writing – review & editing, Investigation, Funding acquisition. Miina Ollikainen: Writing – review & editing, Supervision, Funding acquisition. Sylvain Sebert: Writing – review & editing, Supervision. Elina Sillanpää: Writing – review & editing, Validation, Conceptualization, Supervision, Funding Acquisition.

## Supporting information

eTable 1

eTable 2

## Acknowledgments

We are grateful to all participating twins, their families and teachers, personnel at the Finnish Twin Cohort and researchers at FIMM, who made this study happen. We also thank our collaborators from the Equal Life project for their expertise and dedicated work in collecting, harmonizing, and pre-processing some of the exposome data used in this study - A. Ambros, M. Foraster, J. Julvez, I. van Kamp and B. Raimbault.

## Data availability

The FinnTwin12 data are not publicly available due to the restrictions of informed consent. The FinnTwin12 data are, however, available through the Institute for Molecular Medicine Finland (FIMM) Data Access Committee (DAC) for authorized researchers who have IRB/ethics approval and an institutionally approved study plan. To ensure the protection of privacy and compliance with national data protection legislation, a data use/transfer agreement is needed, the content and specific clauses of which will depend on the nature of the requested data.

