## Supplement for "Exploring the exposome and unexplained variance in biological ageing – insights from a longitudinal twin study in adolescence and early adulthood"

#### Additional description of exposures

##### eTable 1 – Description of exposures

eTable 1 lists all exposures available for specific time points/ages, their sources, units and a short description at the beginning of the study, i.e. before MICE imputation. Variables marked red were removed prior to the analyses due to high missingness (>30%).

*Note: This table is available as a separate Excel file.*

#### Details on missing data

##### eFigure 1 – distribution of missingness

**(A)**

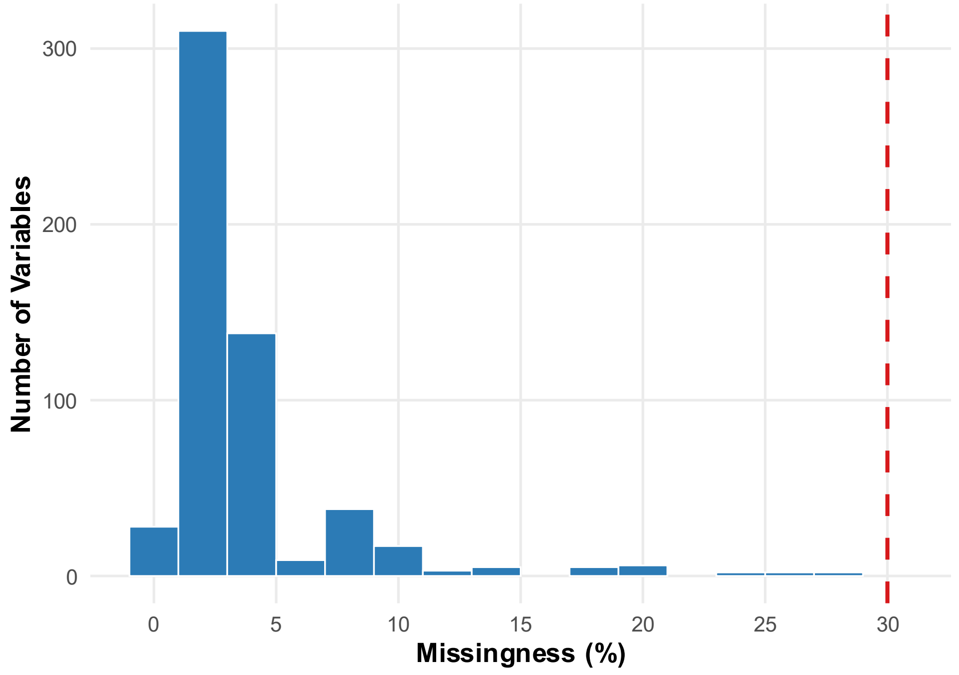

(B)

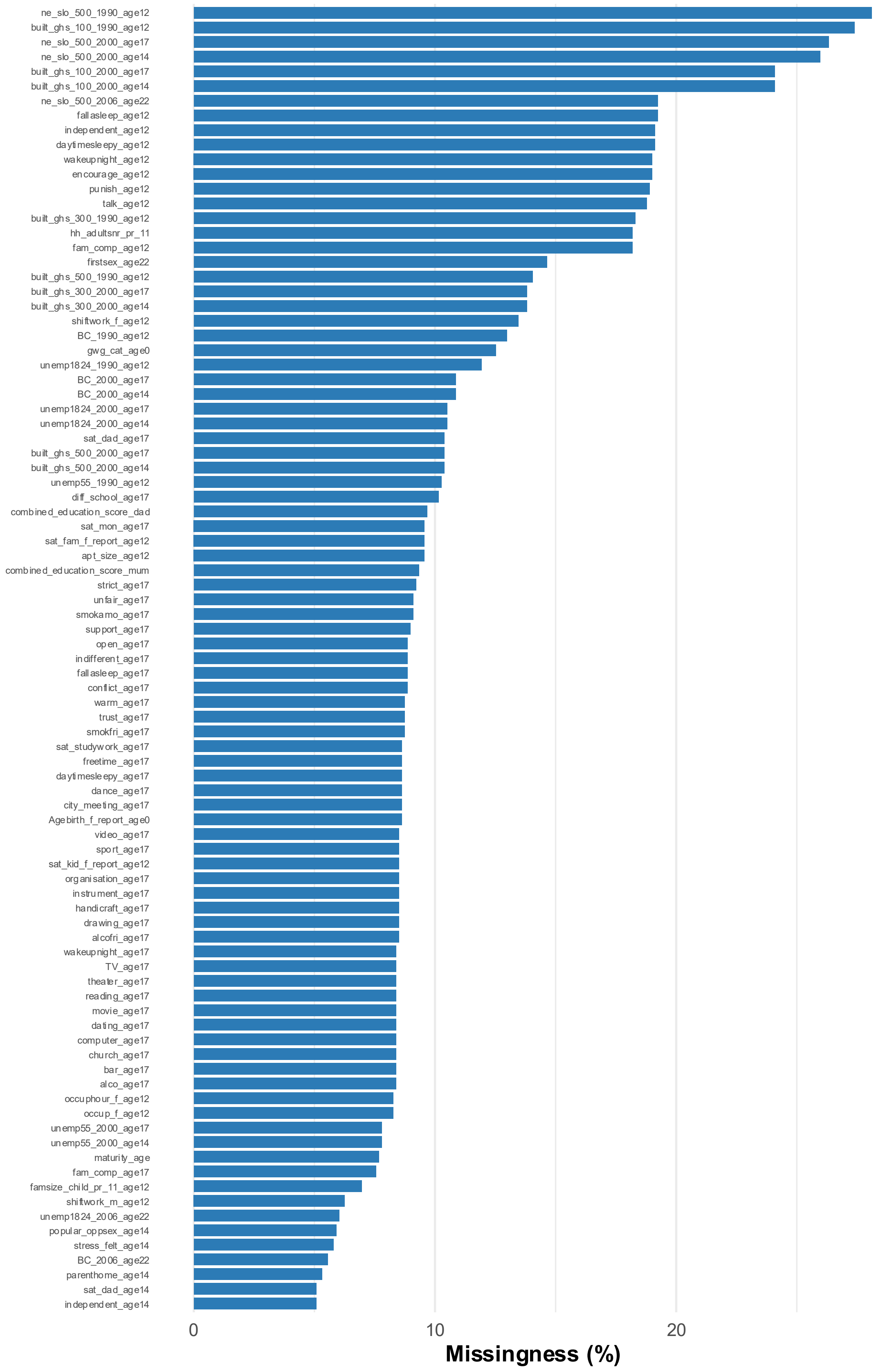

ne_slo_500_1990_age12
built_ghs_100_1990_age12
ne_slo_500_2000_age17
ne_slo_500_2000_age14
built_ghs_100_2000_age17
built_ghs_100_2000_age14
ne_slo_500_2006_age22
fallasleep_age12
independent_age12
daytimesleepy_age12
wakeupnight_age12
encourage_age12
punish_age12
talk_age12
built_ghs_300_1990_age12
hh_adultsnr_pr_11
fam_comp_age12
firstsex_age22
built_ghs_500_1990_age12
built_ghs_300_2000_age17
built_ghs_300_2000_age14
shiftwork_f_age12
BC_1990_age12
gwg_cat_age0
unemp1824_1990_age12
BC_2000_age17
BC_2000_age14
unemp1824_2000_age17
unemp1824_2000_age14
sat_dad_age17
built_ghs_500_2000_age17
built_ghs_500_2000_age14
unemp55_1990_age12
diff_school_age17
combined_education_score_dad
sat_mon_age17
sat_fam_f_report_age12
apt_size_age12
combined_education_score_mum
strict_age17
unfair_age17
smokamo_age17
support_age17
open_age17
indifferent_age17
fallasleep_age17
conflict_age17
warm_age17
trust_age17
smokfri_age17
sat_studywork_age17
freetime_age17
daytimesleepy_age17
dance_age17
city_meeting_age17
Agebirth_f_report_age0
video_age17
sport_age17
sat_kid_f_report_age12
organisation_age17
instrument_age17
handicraft_age17
drawing_age17
alcofri_age17
wakeupnight_age17
TV_age17
theater_age17
reading_age17
movie_age17
dating_age17
computer_age17
church_age17
bar_age17
alco_age17
occuphour_f_age12
occup_f_age12
unemp55_2000_age17
unemp55_2000_age14
maturity_age
fam_comp_age17
famsize_child_pr_11_age12
shiftwork_m_age12
unemp1824_2006_age22
popular_oppsex_age14
stress_felt_age14
BC_2006_age22
parenthome_age14
sat_dad_age14
independent_age14
0
10
20
Missingness (%)

eFigure 1 shows the distribution of missingness across exposome variables.

**(A)** Histogram displaying the percentage of missing values for all exposure variables (N = 565) prior to exclusion. The red dashed line indicates the pre-defined exclusion threshold of 30%. Variables with missingness exceeding this threshold were removed from subsequent analyses to minimize imputation uncertainty. **(B)** Detail of the "imputation zone," showing only variables with missingness between 5% and 30%. These variables were retained but required statistical imputation. Variables with <5% missingness (not shown) were also imputed but carry a negligible risk of bias.

#### eTable 2 - missingness statistics for retained exposure variables

eTable 2 displays missingness counts (N) and percentage (%) prior to imputation alongside observed and imputed statistics (Mean ± SD for continuous, average % of participants for categorical variables) derived from five MICE iterations. Exposures with >30 missingness were excluded beforehand and are not listed here. Imputation was performed on original variables prior to one-hot encoding to preserve categorical structures.

*Note: This table is available as a separate Excel file.*

#### eFigure 2 - Convergence trace plots for representative exposure variables.

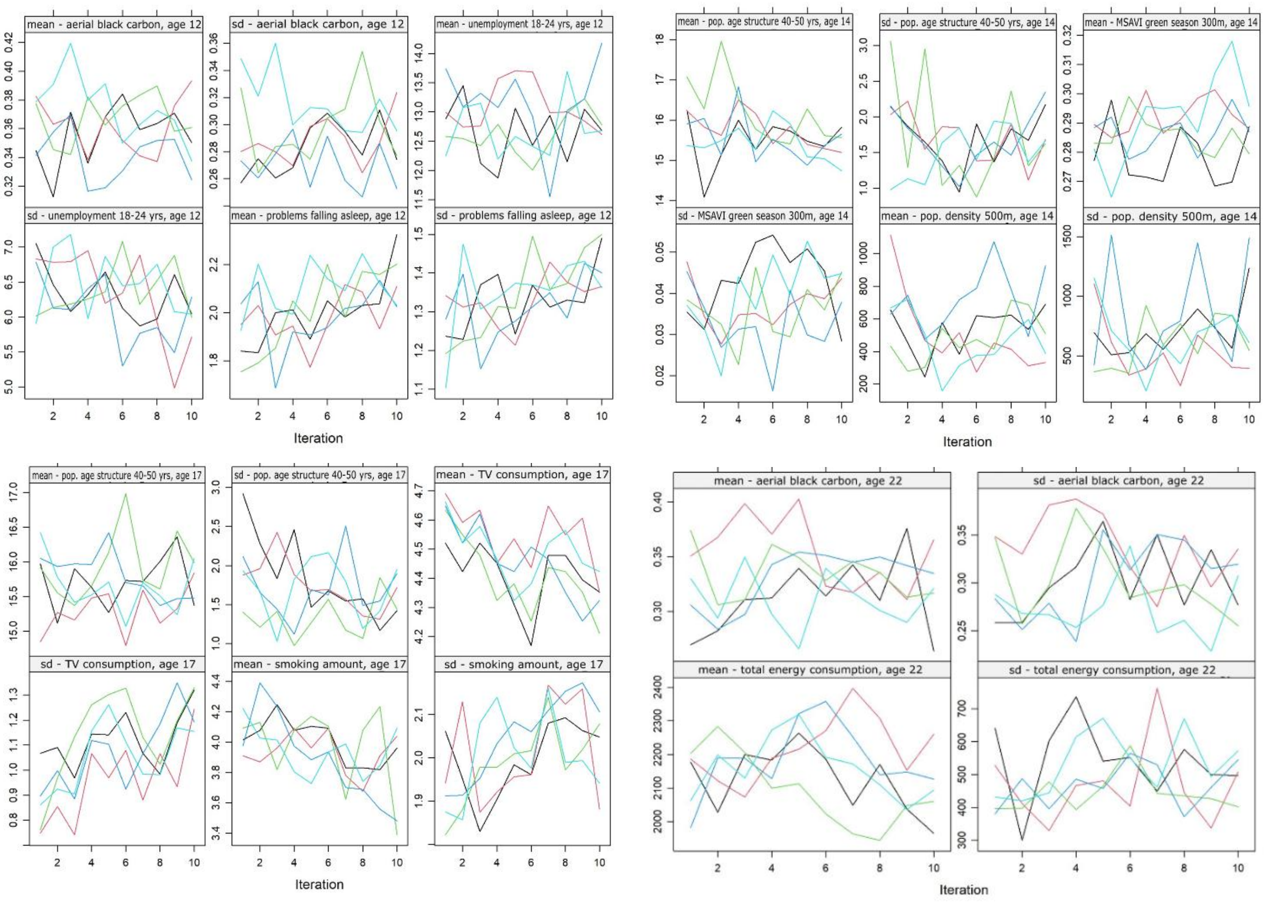

eFigure 2 displays the iteration history for a random selection of exposures in each age class to assess the stability of the MICE algorithm. The x-axis represents the iteration number (1–10), and the y-axis shows the mean of the imputed values. Each coloured line corresponds to one of the five independent imputed datasets. The intermingling of chains without distinct separation or directional trends indicates that the algorithm converged successfully.

#### eFigure 3 – Density plots comparing observed and imputed data distributions for representative exposure variables

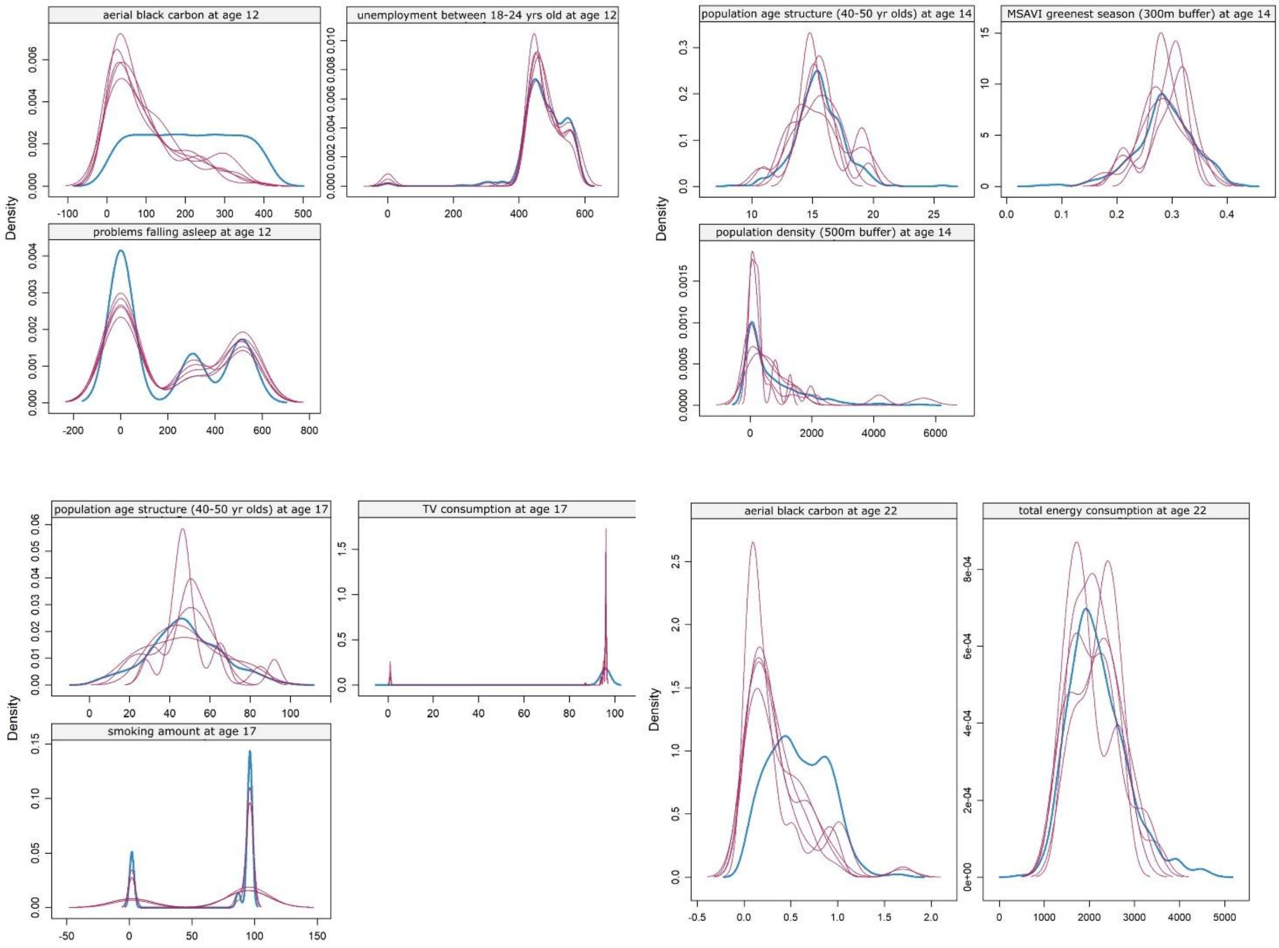

eFigure 3 displays Kernel density estimates for a random selection of exposures in each age class to assess the plausibility of MICE imputed values. The blue line represents the distribution of the originally observed (non-missing) data. The red lines represent the distributions generated in the 5 independent imputed datasets. The strong overlap between the observed and imputed densities indicates that the MICE algorithm preserved the underlying distributional structures.

#### eFigure 4 - Sensitivity analysis comparing effect estimates from complete case vs. imputed data analyses

**(A)**

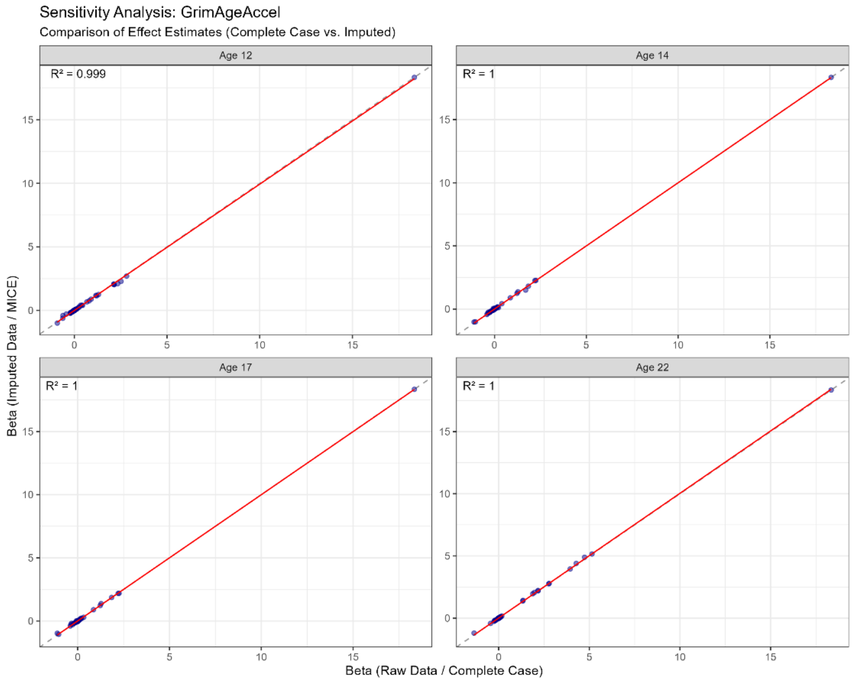

**(B)**

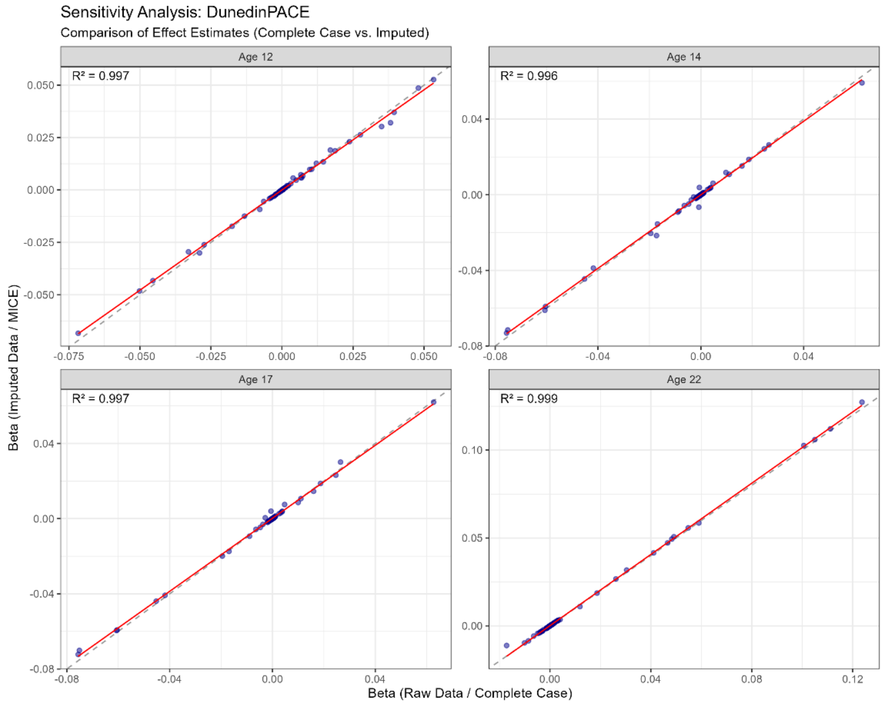

eFigure 4 displays the correlations between regression coefficients calculated using the raw, non-imputed data (complete case analysis, x-axis) and the pooled results from the 5 imputed MICE datasets (y-axis). Each point represents the effect estimate for a single exposure variable on the biological ageing outcome (**(A)** PCGrimAge and **(B)** DunedinPACE), stratified by age group. The dashed diagonal line indicates perfect agreement, while the solid red line shows the linear fit. High coefficients of determination ($R^2$) and strong adherence to the diagonal line demonstrate that the imputation process preserved the underlying exposure-outcome associations and did not introduce systematic bias compared to the complete case analysis.

#### Statistical analyses

##### eTable 3 - ExWAS results for $EA_{GrimAge}$

eTable 3 shows all significant (FDR adjusted p<0.05; in bold) and nominally significant exposures (unadjusted p<0.05) exposures from the ExWAS analysis. Results are listed separately for each age and adjusted for chronological age, sex, zygosity, maternal age at birth and parental education; Parental education level represents the combination score, where higher value=higher overall education. β-estimates represent the average change in $EA_{GrimAge}$ per 1-category increase.

| Age class | Exposure | Exposure Family | Levels/ Units | Estimate β (95 % CI) | unadj. p-value | p- value (FDR-adj.) |
|---|---|---|---|---|---|---|
| **age 12** | **sex (male)** | **covariate** | **male-female** | **1.27 (0.7, 1.9)** | **<0.0001** | **0.002** |
| | **paternal education level** | **covariate** | **1-9** | **-0.36 (-0.64, -0.07)** | **0.016** | **0.53** |
| | voluntary activity in organisations | lifestyle | 1 daily<br>2 a few times a week<br>3 a few times a month<br>4 a few times in 6 months<br>5 never | -1.08 (-0.64, -0.07) | 0.016 | 0.53 |
| | parental appreciation of offspring opinions | family and parents | 1 almost always<br>2 generally<br>3 only sometimes<br>4 seldom or never | -0.44 (-0.86, -0.02) | 0.041 | 0.788 |
| | family composition | family and parents | 1 the twins' biological father<br>2 the twins' biological mother<br>3 siblings<br>4 other adults (new partner, grandparents, other relatives, others)<br>5 other children (halfsiblings, adopted siblings) | 0.29 (0.03, 0.54) | 0.026 | 0.708 |
| | maternal working hours | family and parents | h/d | 0.34 (0.09, 0.59) | 0.008 | 0.509 |
| | smoking during pregnancy | lifestyle | 1 I smoked on average x cigarettes/day<br>2 I quit smoking in pregnancy month x<br>3 I have never smoked | 0.27 (0.01, 0.52) | 0.041 | 0.788 |
| **age 14** | **sex (male)** | **covariate** | **male-female** | **1.14 (0.52, 1.75)** | **0.0003** | **0.049** |
| | **paternal education level** | **covariate** | **1-9** | **-0.36 (-0.65, -0.06)** | **0.017** | **0.661** |
| | physical activity indoors | lifestyle | 1 almost daily<br>2 more than once a week<br>3 about once a week<br>4 about once a month<br>5 seldom or never | 0.73 (0.09, 1.36) | 0.025 | 0.661 |
| | daytime sleepiness | lifestyle | 1 almost daily<br>2 more than once a week<br>3 about once a week<br>4 about once a month<br>5 seldom or never | 0.8 (0.08, 1.52) | 0.03 | 0.661 |
| | pocket money use agreements | family and parents | 1 almost always<br>2 generally<br>3 only sometimes<br>4 seldom or never | -0.51 (-1.00, -0.02) | 0.041 | 0.826 |
| | stress due to parental separation/divorce | stressful life event | yes/no | 1.21 (0.26, 2.15) | 0.013 | 0.661 |
| | treecover | living environment | 100m buffer | -0.31 (-0.57, -0.05) | 0.018 | 0.661 |
| | residential area: population age structure (40-50 yrs) | demographics | % of population | -0.28 (-0.53, -0.03) | 0.031 | 0.661 |

| | | | | | | |
|---|---|---|---|---|---|---|
| | residential area: foreign citizens | demographics | % of population | 0.25 (0.00, 0.5) | 0.049 | 0.661 |
| **age 17** | **sex (male)** | **covariate** | **male-female** | **1.22 (0.78, 1.65)** | **<0.001** | **<0.001** |
| | **maternal education level** | **covariate** | **1-9** | **-0.32 (-0.63, -0.02)** | **0.038** | **0.545** |
| | **paternal education level** | **covariate** | **1-9** | **-0.30 (-0.59, -0.02)** | **0.037** | **0.545** |
| | **amount of smoking** | **lifestyle** | **1 daily<br>2 at least once a week<br>3 less than once a week<br>4 trying to or have quit<br>5 never** | **-1.63 (-2.55, -0.71)** | **0.001** | **0.040** |
| | voluntary activity in organisations | lifestyle | 1 daily<br>2 a few times a week<br>3 a few times a month<br>4 a few times in 6 months<br>5 never | -0.75 (-1.32, -0.18) | 0.010 | 0.269 |
| | family composition | family and parents | 1 biological father<br>2 biological mother<br>3 siblings<br>4 other adults (new partner, grandparents<br>5 other children (halfsiblings, adopted siblings) | 0.36 (0.11, 0.61) | 0.004 | 0.140 |
| | amount of sport activities | lifestyle | 1 not at all<br>1 less than once a month<br>3 1-2 times a month<br>4 about once a week<br>5 2-3 times a week<br>6 4-5 times a week<br>7 just about every day | -2.00 (-3.32, -0.69) | 0.003 | 0.123 |
| | tree cover | living environment | % | -0.28 (-0.53, -0.02) | 0.023 | 0.527 |
| | residential area: population age structure (40-50 yrs) | demographics | % of total population | -0.28 (-0.52, -0.03) | 0.029 | 0.527 |
| **age 22** | **sex (male)** | **covariate** | **male-female** | **-0.01 (-0.01, -0.00)** | **<0.001** | **<0.001** |
| | **paternal education level** | **covariate** | **1-9** | **-0.4 (-0.69, -0.11)** | **0.008** | **0.098** |
| | **amount of smoking** | **lifestyle** | **1 daily<br>2 at least once a week<br>3 less than once a week<br>4 trying to or have quit<br>5 never** | **-1.77 (-2.57, -0.97)** | **<0.001** | **0.005** |
| | **amount of alcohol consumption** | **lifestyle** | **g/d** | **0.60 (0.39, 0.81)** | **<0.001** | **<0.001** |
| | **number of sex partners** | **lifestyle** | **1 I have not had sex<br>2 one<br>3 two<br>4 three or four<br>5 five or more** | **0.51 (0.31, 0.72)** | **<0.001** | **0.001** |
| | **age at sexual debut** | **lifestyle** | **age in years** | **-0.49 (-0.73, -0.25)** | **0.0001** | **0.003** |
| | **residential area: crime rate (crimes against life and death) at age 22** | **demographics** | **per 1000 population** | **-0.36 (-0.58, -0.13)** | **0.002** | **0.037** |
| | total energy intake | lifestyle | kcal/d | 0.34 (0.11, 0.57) | 0.003 | 0.054 |
| | total fat intake | lifestyle | g/d | 0.23 (0.00, 0.45) | 0.047 | 0.281 |
| | total sugar intake | lifestyle | g/d | 0.23 (0.03, 0.44) | 0.024 | 0.202 |
| | area of closest blue space | living environment | squaremeter | -0.26 (-0.48, -0.04) | 0.023 | 0.202 |
| | vegetation index during greenest season (MSAVI) | living environment | 500m buffer | 0.23 (0.01, 0.44) | 0.043 | 0.281 |
| | aerial black carbon | living environment | ppm | -0.28 (-0.52, -0.05) | 0.018 | 0.193 |
| | residential area: education level (only primary education) | demographics | % of population | 0.31 (0.08, 0.54) | 0.008 | 0.098 |
| | residential area: unemployment rate (18-24 year olds) | demographics | % of population | 0.24 (0.01, 0.48) | 0.043 | 0.281 |
| | residential area: unemployment rate (women) | demographics | % of population | 0.30 (0.06, 0.53) | 0.013 | 0.148 |

#### eTable 4 - ExWAS results for $EA_{DunedinPACE}$

eTable 3 shows all significant (FDR adjusted p<0.05; in bold) and nominally significant exposures (unadjusted p<0.05) exposures from the ExWAS analysis. Results are listed separately for each age and adjusted for chronological age, sex, zygosity, maternal age at birth and parental education; Parental education level represents the combination score, where higher value=higher overall education. β-estimates represent the average change in $EA_{DunedinPACE}$ per 1-category increase.

| Age class | Exposure | Exposure Family | Levels/ Units | Estimate β (95 % CI) | unadj. p-value | p- value (FDR-adjusted) |
|---|---|---|---|---|---|---|
| **age 12** | **sex (male)** | **covariate** | **male-female** | **-0.05 (-0.06, -0.03)** | **<0.001** | **<0.001** |
| | **paternal education level** | **covariate** | **1-9** | **-0.01 (-0.02, -0.00)** | **0.017** | **0.577** |
| | maternal working hours | family and parents | h/d | 0.01 (0.00, 0.02) | 0.004 | 0.226 |
| | maternal shiftwork | family and parents | 1 regular day work<br>2 regular evening work<br>3 regular night work<br>4 irregular work<br>5 shiftwork without nights<br>6 shiftwork + nights<br>7 three-shift work | 0.01 (0.00, 0.02) | 0.023 | 0.609 |
| | apartment type | living environment | 1 detached house<br>2 semi-detached house<br>3 terraced house<br>4 apartment house<br>5 other | 0.01 (0.00, 0.02) | 0.005 | 0.226 |
| | residential area: population count | demographics | 300m buffer | 0.009 (0.00, 0.02) | 0.035 | 0.790 |
| | residential area: household structure (singles) | demographics | % of population | 0.01 (0.00, 0.02) | 0.046 | 0.860 |
| **age 14** | **sex (male)** | **covariate** | **male-female** | **-0.05 (-0.06, -0.03)** | **<0.001** | **<0.001** |
| | **paternal education level** | **covariate** | **1-9** | **-0.01 (-0.02, -0.00)** | **0.019** | **0.509** |
| | daytime sleepiness | lifestyle | 1 almost daily<br>2 more than once a week<br>3 about once a week<br>4 about once a month<br>5 seldom or never | 0.03 (0.00, 0.05) | 0.020 | 0.509 |
| | sport activities together with parents | family and parents | 1 daily<br>2 two times/week<br>3 two times/month<br>4 two times/half a year<br>5 never | 0.04 (0.00, 0.07) | 0.042 | 0.579 |
| | stress due to change of residence | stressful life event | yes/no | 0.02 (0.00, 0.05) | 0.044 | 0.579 |
| | stress due to parent's new spouse moving in | stressful life event | yes/no | 0.04 (0.00, 0.08) | 0.047 | 0.579 |
| | treecover | living environment | 100m buffer | -0.01 (-0.02, -0.01) | 0.001 | 0.085 |
| | treecover | living environment | 300m buffer | -0.01 (-0.02, -0.00) | 0.014 | 0.509 |
| | residential area: population age structure (40-50 yrs) | demographics | % of population | -0.01 (-0002, -0.00) | 0.022 | 0.509 |
| | residential area: population count | demographics | 300m buffer | 0.01 (0.00, 0.02) | 0.015 | 0.509 |
| | residential area: population count | demographics | 500m buffer | 0.01 (0.00, 0.02) | 0.025 | 0.509 |
| **age 17** | **sex (male)** | **covariate** | **male-female** | **-0.05 (-0.06, -0.03)** | **<0.001** | **<0.001** |
| | **paternal education level** | **covariate** | **1-9** | **-0.01 (-0.02, <0.00)** | **0.046** | **0.660** |
| | family composition | family and parents | 1 biological father<br>2 biological mother<br>3 siblings<br>4 other adults (new partner, grandparents<br>5 other children (halfsiblings, adopted siblings) | 0.01 (0.00, 0.02) | 0.003 | 0.113 |

| | | | | | | |
|---|---|---|---|---|---|---|
| | residential area: population count | demographics | 300m buffer | 0.01 (0.00, 0.02) | 0.016 | 0.423 |
| | residential area: population count | demographics | 500m buffer | 0.01 (0.00, 0.02) | 0.029 | 0.513 |
| | residential area: built-up environment | living environment | 300m buffer | 0.01 (0.00, 0.02) | 0.037 | 0.595 |
| | tree cover | living environment | 100m buffer | -0.01 (-0.02, -0.01) | 0.002 | 0.113 |
| | tree cover | living environment | 300m buffer | -0.01 (-0.02, -0.00) | 0.017 | 0.421 |
| | population age structure at residence (40-50 year olds) | demographics | % of population | -0.01 (-0.02, -0.00) | 0.020 | 0.421 |
| **age 22** | **sex (male)** | **covariate** | **male-female** | **-0.05 (-0.06, -0.03)** | **<0.001** | **<0.001** |
| | **paternal education level** | **covariate** | **1-9** | **-0.01 (-0.02, -0.00)** | **0.023** | **0.281** |
| | **amount of alcohol** | **lifestyle** | **g/d** | **0.01 (0.01, 0.02)** | **<0.001** | **0.018** |
| | **area of closest green space** | **living environment** | **squaremeter** | **0.01 (0.01, 0.02)** | **<0.001** | **0.010** |
| | amount of smoking | lifestyle | 1 daily<br>2 at least once a week<br>3 less than once a week<br>4 trying to or have quit<br>5 never | -0.04 (-0.07, -0.01) | 0.003 | 0.093 |
| | age at sexual debut | lifestyle | age in years | -0.01 (-0.02, -0.00) | 0.014 | 0.267 |
| | number of sex partners | lifestyle | 1 I have not had sex<br>2 one<br>3 two<br>4 three or four<br>5 five or more | 0.01 (0.00, 0.16) | 0.005 | 0.131 |
| | total fibre intake | lifestyle | g/d | -0.01 (-0.02, -0.00) | 0.015 | 0.267 |
| | vegetation index during greenest season (NDVI) | living environment | 500m buffer | 0.01 (0.00, 0.01) | 0.040 | 0.281 |
| | vegetation index during greenest season (MSAVI) | living environment | 300m buffer | 0.01 (0.00, 0.01) | 0.034 | 0.281 |
| | vegetation index during greenest season (MSAVI) | living environment | 500m buffer | 0.01 (0.00, 0.02) | 0.021 | 0.281 |
| | vegetation index during all seasons (MSAVI) | living environment | 300m buffer | 0.01 (<0.00, 0.01) | 0.049 | 0.281 |
| | vegetation index during all seasons (MSAVI) | living environment | 500m buffer | 0.01 (<0.00, 0.01) | 0.048 | 0.281 |
| | crime rate in living area (crimes against life and health) | living environment | offences per 1000 population | -0.01 (-0.02, -0.00) | 0.034 | 0.281 |
| | population age structure at residence (20-30 yrs) | demographics | % of population | -0.01 (-0.01, -0.00) | 0.044 | 0.281 |
| | population age structure at residence (60-65 yrs) | demographics | % of population | 0.01 (0.00, 0.02) | 0.033 | 0.281 |
| | population age structure at residence (>66 yrs) | demographics | % of population | 0.01 (<0.00, 0.01) | 0.050 | 0.281 |
| | residential area: education level (only primary education) | demographics | % of population | 0.01 (0.00, 0.02) | 0.020 | 0.281 |
| | residential area: income (second lowest quartile) | demographics | % of households | 0.01 (0.00, 0.02) | 0.029 | 0.281 |
| | residential area: unemployment rate (total) | demographics | % of population | 0.01 (<0.00, 0.02) | 0.049 | 0.281 |
| | residential unemployment rate (women) | demographics | % of population | 0.01 (0.00, 0.02) | 0.032 | 0.281 |

#### eTable 5 – stable predictors identified by the Knock-off boosted tree (KOBT) analysis

eTable 5 lists predictors selected by the Knockoff Boosted Tree (KOBT) algorithm to control the False Discovery Rate (FDR) at 0.20. The analysis was performed across five multiple imputed datasets, and variables were retained if they were selected in > 50% of the imputations (stability selection). "Shared" indicates predictors identified as significant in both analysis modes. "Unique to Longitudinal" indicates predictors identified only when repeated measures were grouped to represent lifelong exposure. "Unique to Cross-Sectional" indicates predictors identified only when specific ages were treated as independent features.

| Outcome | Mode | selected exposures |
|---|---|---|
| GrimAge | Longitudinal and Cross-sectional | residential area: education level (post-secondary non-tertiary) at age 22 |
| | | population density (500m buffer) at age 17 |
| | | smoking status at age 17 |
| | | total alcohol consumption at age 22 |
| | | residential area: unemployment rate (18-24 yrs old) at age 22 |
| GrimAge | Longitudinal | residential area: household structure (married couples without underaged children) at age 14 |
| | | residential area: household structure (married couples without underaged children) at age 17 |
| | | population density (500m buffer) at age 14 |
| | | smoking status at age 14 |
| GrimAge | Cross-Sectional | vegetation index (MSAVI) during greenest season (300m buffer) at age 14 |
| | | vegetation index (NDVI) during all seasons (100m buffer) at age 12 |
| DunedinPACE | Longitudinal and Cross-sectional | maternal age at birth |
| | | size of residence at age 12 |
| | | paternal education score |
| | | female gender |
| | | residential area: income level (second lowest) at age 22 |
| | | vegetation index (MSAVI) during greenest season (300m buffer) at age 14 |
| | | satisfaction with school at age 12 |
| | | smoking status at age 17 |
| | | smoking status at age 22 |
| | | treecover (100m buffer) at age 17 |
| DunedinPACE | Longitudinal | vegetation index (MSAVI) during greenest season (300m buffer) at age 17 |
| | | satisaction with school at age 14 |
| | | smoking status at age 14 |
| | | treecover (100m buffer) at age 14 |
| DunedinPACE | Cross-Sectional | None |

#### eFigure 5 – KOBT stability selection frequency of predictors for $EA_{GrimAge}$ and $EA_{DunedinPACE}$ across multiple imputations

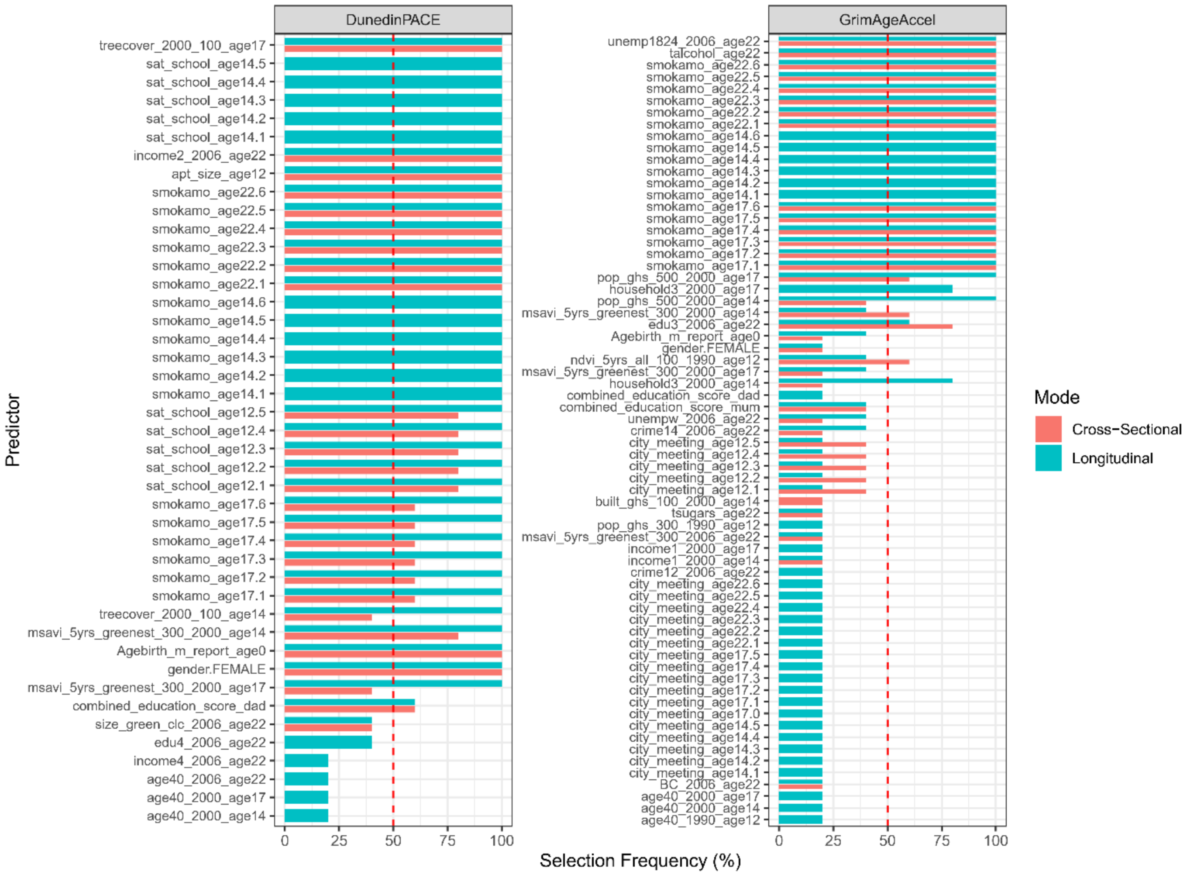

eFigure 5: Bar lengths represent the stability score (percentage of imputed datasets in which a specific predictor was selected by the KOBT algorithm) for each variable. Results are stratified by analysis mode: Longitudinal (grouping repeated measures) and Cross-Sectional (treating time points independently). The dashed red line represents the stability threshold of 50%; only variables exceeding this threshold are considered robustly selected. Variables with higher stability scores demonstrate stronger evidence of association with the aging biomarkers, robust to missing data uncertainty.

#### eTable 6 – Results of sparse N-way Partial Least Squares (sNPLS) analysis

eTable 6: The sNPLS algorithm was applied to the three-dimensional data array (Individual × Exposure × Time) to identify stable linear predictors of $EA_{GrimAge}$ and $EA_{DunedinPACE}$. Cross-validation was performed within each imputed dataset. "None Selected" indicates that no variables met the stability threshold, highlighting the absence of strong linear multidimensional signals.

| outcome | predictive exposure | result |
|---|---|---|
| PCGrimAge | none selected | no stable predictors identified |
| DunedinPACE | none selected | no stable predictors identified |

#### eTable 7 – Exposures predictive of $EA_{DunedinPACE}$ selected by the Boruta algorithm

eTable 7 lists all $EA_{DunedinPACE}$ predictive exposures selected by the Boruta Stability Selection algorithm, including their model performance importance score (%IncMSE) and ranked by absolute mean SHAP values.

| exposure name | exposure | Absolute Mean SHAP value | Importance score (%IncMSE) |
|---|---|---|---|
| gender.FEMALE | female gender | 0.012339306 | 17.417806 |
| gender.MALE | male gender | 0.010797007 | 15.323339 |
| treecover_2000_100_age14 | treecover (100m buffer) at age 14 | 0.009860335 | 16.376558 |
| apt_size_age12 | size of residence at age 12 | 0.009233769 | 19.652088 |
| treecover_2000_100_age17 | treecover in % (100m buffer) at age 17 | 0.007161865 | 15.004908 |
| smokamo_age22.1 | daily smoking at age 22 | 0.006926661 | 11.368704 |
| Agebirth_m_report_age0 | maternal age at birth | 0.00619897 | 15.739127 |
| size_green_clc_2006_age22 | area of closest green space at age 22 | 0.00543822 | 15.094225 |
| built_ghs_100_2000_age14 | built environment (100m buffer) at age 14 | 0.005017659 | 14.077984 |
| msavi_5yrs_greenest_300_2000_age14 | MSAVI in greenest season (300m buffer) at age 14 | 0.004454121 | 13.476106 |
| age40_2000_age17 | population age structure (40-50 yrs) at age 17 | 0.004370229 | 13.954822 |
| age40_2000_age14 | population age structure (40-50 yrs) at age 14 | 0.00416443 | 13.43333 |
| msavi_5yrs_greenest_300_2000_age17 | MSAVI in greenest season (300m buffer) at age 17 | 0.003882735 | 14.229004 |
| smokamo_age17.1 | daily smoking at age 17 | 0.003873663 | 9.982877 |
| household2_2000_age14 | household structure (singles with underaged children) at age 14 | 0.003500827 | 12.493706 |
| household2_2000_age17 | household structure (singles with underaged children) at age 17 | 0.003087508 | 13.261387 |
| treecover_2000_500_age14 | treecover_2000_500_age14 | 0.003050642 | 11.437823 |
| treecover_2000_300_age17 | treecover in % (300m buffer) at age 17 | 0.002955807 | 9.338284 |
| treecover_2000_300_age14 | treecover in % (300m buffer) at age 14 | 0.002836949 | 12.499407 |
| ndvi_5yrs_greenest_500_2000_age14 | ndvi_5yrs_greenest_500_2000_age14 | 0.002789229 | 14.401908 |
| msavi_5yrs_greenest_100_2000_age14 | MSAVI in greenest season (100m buffer) at age 14 | 0.002687632 | 10.800851 |
| ndvi_5yrs_greenest_500_2000_age17 | NDVI in greenest season (500m buffer) at age 17 | 0.002590753 | 13.74587 |
| ndvi_5yrs_all_300_2000_age17 | ndvi_5yrs_all_300_2000_age17 | 0.002574914 | 11.94731 |
| ndvi_5yrs_greenest_300_2000_age14 | ndvi_5yrs_greenest_300_2000_age14 | 0.002562321 | 14.068048 |
| ndvi_5yrs_greenest_100_2000_age14 | ndvi_5yrs_greenest_100_2000_age14 | 0.002466814 | 12.584507 |
| pop_ghs_500_2000_age17 | population count (500m buffer) at age 17 | 0.002461175 | 12.053548 |
| msavi_5yrs_greenest_100_2000_age17 | MSAVI in greenest season (100m buffer) at age 17 | 0.002452825 | 10.110715 |
| msavi_5yrs_all_300_2000_age14 | msavi_5yrs_all_300_2000_age14 | 0.002436468 | 12.107696 |
| pop_ghs_500_2000_age14 | population count (500m buffer) at age 14 | 0.002425415 | 10.208247 |
| ndvi_5yrs_all_100_2000_age17 | ndvi_5yrs_all_100_2000_age17 | 0.002403084 | 9.628024 |
| ndvi_5yrs_all_300_2000_age14 | ndvi_5yrs_all_300_2000_age14 | 0.002378795 | 12.072979 |
| msavi_5yrs_all_500_2000_age17 | MSAVI in all seasons (500m buffer) at age 17 | 0.002299382 | 10.455301 |
| pop_ghs_300_2000_age17 | pop_ghs_300_2000_age17 | 0.002273326 | 12.490133 |
| msavi_5yrs_all_100_2000_age14 | msavi_5yrs_all_100_2000_age14 | 0.002213585 | 11.799457 |
| msavi_5yrs_greenest_500_2000_age14 | msavi_5yrs_greenest_500_2000_age14 | 0.002204507 | 12.717323 |
| pop_ghs_300_2000_age14 | population count (300m buffer) at age 14 | 0.002087277 | 11.656613 |
| msavi_5yrs_all_500_2000_age14 | msavi_5yrs_all_500_2000_age14 | 0.002063022 | 13.036406 |
| ndvi_5yrs_greenest_300_2000_age17 | ndvi_5yrs_greenest_300_2000_age17 | 0.002058805 | 11.845796 |
| ndvi_5yrs_greenest_100_2000_age17 | ndvi_5yrs_greenest_100_2000_age17 | 0.001939078 | 10.195133 |

#### eTable 8 - Comparison of model performance between the full set of exposures and the Boruta-selected predictive exposures

eTable 8 compares the predictive performance (Out-of-Bag $R^2$) of a Random Forest model trained on the full exposure set (n >500 exposures) versus the final selected model trained only on features confirmed relevant by the Boruta algorithm.

| outcome | baseline model ($R^2$) | selected model ($R^2$) |
|---|---|---|
| PCGrimAge | 25.70% | 18.60% |
| DunedinPACE | 30.80% | 22.40% |

#### eTable 9 - Results of Linear Mixed-Effects Models (LMM) for $EA_{Grimage}$

eTable 9 presents pooled fixed-effect estimates (β) and 95% confidence intervals across five imputed datasets for $EA_{Grimage}$. Predictors were standardized (Z-score) prior to analysis. Random intercepts were included for *familyid* to account for the nested structure of the twin data. Models were adjusted for all Boruta-selected predictive exposures simultaneously. Significance threshold p≤ 0.05.

| | estimate [95% CI] | std.error | statistic | df | p-value |
|---|---|---|---|---|---|
| aerial black carbon at age 22 | -0.29 [-0.55, -0.02] | 0.13618128 | -2.1006103 | 464.947 | 0.03621 |
| age at sexual debut | -0.11 [-0.33, 0.12] | 0.11321282 | -0.9446124 | 141.187 | 0.34647 |
| vegetation index (MSAVI) during greenest season (100m buffer) at age 14 | -0.49 [-2.60, 1.63] | 1.00196273 | -0.4851884 | 16.6579 | 0.63386 |
| vegetation index (MSAVI) during greenest season (100m buffer) at age 17 | 0.14 [-1.94, 2.23] | 0.98593009 | 0.14697864 | 16.9841 | 0.88488 |
| daily smoking at age 17 | 0.48 [-0.11, 1.07] | 0.30083417 | 1.59007686 | 219.256 | 0.11326 |
| daily smoking at age 22 | 2.24 [1.69, 2.78] | 0.27661043 | 8.08226802 | 768.716 | 2.46E-15 |
| no smoking at age 22 | -0.19 [-0.63, 0.25] | 0.22269458 | -0.8451713 | 827.838 | 0.39826 |
| total alcohol consumption at age 22 | 0.37 [0.16, 0.58] | 0.1066878 | 3.50370735 | 449.753 | 0.0005 |
| total energy consumption at age 22 | 0.23 [0.02, 0.44] | 0.10514388 | 2.17557005 | 471.247 | 0.03008 |
| vegetation index (MSAVI) during greenest season (300m buffer) at age 14 | 0.52 [0.06, 0.98] | 0.23292798 | 2.22988354 | 598.384 | 0.02613 |
| residential area: unemployment rate (18-24 yrs old) at age 22 | 0.24 [-0.01, 0.49] | 0.12905124 | 1.85709236 | 396.574 | 0.06404 |
| residential area: education level (master's degree or higher) at age 22 | -0.11 [-0.35, 0.13] | 0.12322218 | -0.8896034 | 822.758 | 0.37394 |
| population density level (500m buffer) at age 14 | 0.49 [0.19, 0.78] | 0.14992156 | 3.23685584 | 719.906 | 0.00126 |

#### eTable 10 - Results of Linear Mixed-Effects Models (LMM) for $EA_{DunedinPACE}$

eTable 10 presents pooled fixed-effect estimates (β) and 95% confidence intervals across five imputed datasets for $EA_{DunedinPACE}$. Predictors were standardized (Z-score) prior to analysis. Random intercepts were included for *familyid* to account for the nested structure of the twin data. Models were adjusted for all Boruta-selected predictive exposures simultaneously. Significance threshold p≤ 0.05.

| | estimate [95% CI] | std.error | statistic | df | p-value |
|---|---|---|---|---|---|
| size of residence at age 12 | 0.00 [-0.01, 0.00] | 0.003847965 | -1.25001518 | 789.5722049 | 0.211664364 |
| female gender | 0.04 [0.03, 0.06] | 0.006820198 | 6.589338679 | 801.4043606 | 8.00E-11 |
| residential area: household structure (married couples without underaged children) at age 14 | 0.05 [-0.96, 1.06] | 0.422646338 | 0.112793319 | 6.663740245 | 0.913516668 |
| vegetation index (MSAVI) during all seasons (500m buffer) at age 14 | 0.06 [-0.67, 0.79] | 0.362324817 | 0.168613976 | 43.72283468 | 0.8668779 |
| vegetation index (MSAVI) during all seasons (500m buffer) at age 17 | -0.01 [-0.74, 0.72] | 0.364133871 | -0.028786244 | 43.24985421 | 0.977167496 |
| vegetation index (MSAVI) during greenest season (100m buffer) at age 14 | 0.03 [-0.35, 0.40] | 0.176097803 | 0.143167868 | 14.940305 | 0.888070676 |
| vegetation index (MSAVI) during greenest season (300m buffer) at age 14 | 0.14 [-0.71, 1.00] | 0.379163209 | 0.373447745 | 9.308246355 | 0.717179028 |
| vegetation index (MSAVI) during greenest season (300m buffer) at age 17 | -0.12 [-0.97, 0.73] | 0.380954173 | -0.318940491 | 9.757913059 | 0.756493862 |
| vegetation index (NDVI) during all seasons (300m buffer) at age 14 | -0.12 [-0.41, 0.17] | 0.144745345 | -0.806967819 | 56.23846062 | 0.423084768 |
| vegetation index (NDVI) during all seasons (300m buffer) at age 17 | 0.04 [-0.22, 0.30] | 0.130963914 | 0.276209167 | 130.060088 | 0.782825838 |
| vegetation index (NDVI) during greenest season (500m buffer) at age 14 | -0.15 [-0.81, 0.51] | 0.296957766 | -0.510806783 | 10.09884341 | 0.620466424 |
| vegetation index (NDVI) during greenest season (500m buffer) at age 17 | 0.14 [-0.52, 0.81] | 0.298978148 | 0.477674092 | 9.918443399 | 0.64323445 |
| population density (300m buffer) at age 14 | -0.11 [-1.02, 0.79] | 0.373820514 | -0.30462046 | 6.322112274 | 0.770435384 |
| population density (300m buffer) at age 17 | 0.12 [-0.78, 1.02] | 0.372526222 | 0.331184383 | 6.334894176 | 0.751187899 |
| population density (500m buffer) at age 14 | 0.10 [-0.31, 0.50] | 0.193589768 | 0.498237292 | 17.08856517 | 0.624668286 |
| population density (500m buffer) at age 17 | -0.09 [-0.50, 0.31] | 0.192375481 | -0.482599142 | 17.38099548 | 0.635402404 |
| size of closest green space at age 22 | 0.01 [0.01, 0.02] | 0.003417187 | 3.44993484 | 801.7353556 | 0.000590017 |
| daily smoking at age 17 | 0.02 [0.00, 0.04] | 0.009692197 | 1.72323478 | 287.6353051 | 0.085920749 |
| daily smoking at age 22 | 0.04 [0.02, 0.06] | 0.008855336 | 4.315845136 | 538.9418456 | 1.89E-05 |
| treecover (100m buffer) at age 14 | 0.00 [-0.15, 0.15] | 0.068459503 | 0.004759662 | 13.6825589 | 0.996271053 |
| treecover (100m buffer) at age 17 | -0.01 [-0.16, 0.14] | 0.069417648 | -0.154946868 | 13.13895981 | 0.879218205 |
| treecover (300m buffer) at age 14 | 0.00 [-0.29, 0.29] | 0.147640248 | 0.005003149 | 241.2908145 | 0.996012216 |
| treecover (300m buffer) at age 17 | -0.02 [-0.31, 0.27] | 0.147334586 | -0.112631846 | 210.4198209 | 0.910429814 |
| maternal age at birth | 0.00 [0.00, 0.01] | 0.003926187 | 1.159068456 | 773.2809135 | 0.246786075 |
| residential area: age structure (40-50 yrs old) at age 17 | 0.09 [-0.47, 0.65] | 0.246391228 | 0.364452887 | 8.522393396 | 0.724398043 |
| vegetation index (MSAVI) during all seasons (300m buffer) at age 14 | 0.00 [-0.14, 0.15] | 0.073503779 | 0.013884564 | 113.4975252 | 0.988946454 |
| vegetation index (MSAVI) during greenest season (100m buffer) at age 17 | 0.02 [-0.28, 0.31] | 0.148658367 | 0.113557399 | 62.26557193 | 0.909954014 |
| vegetation index (NDVI) during all seasons (100m buffer) at age 17 | 0.07 [-0.16, 0.30] | 0.100194032 | 0.694965075 | 8.364291602 | 0.505921597 |
| vegetation index (NDVI) during greenest season (100m buffer) at age 17 | -0.08 [-0.39, 0.23] | 0.14804133 | -0.557597661 | 18.2737037 | 0.583885906 |
| vegetation index (NDVI) during greenest season (300m buffer) at age 14 | 0.11 [-0.77, 0.99] | 0.371921949 | 0.295907465 | 7.029840284 | 0.775850777 |
| vegetation index (NDVI) during greenest season (300m buffer) at age 17 | -0.04 [-0.92, 0.84] | 0.378317523 | -0.1028923 | 7.586500317 | 0.92071594 |
| residential area: age structure (40-50 yrs old) at age 14 | -0.10 [-0.66, 0.47] | 0.246406582 | -0.393781803 | 8.534085484 | 0.703399087 |
| built environment (100m buffer) at age 14 | 0.00 [-0.02, 0.01] | 0.00656753 | -0.365191277 | 175.3238224 | 0.715408802 |
| residential area: household structure (single parents with underaged children) at age 17 | -0.04 [-1.05, 0.97] | 0.4221757 | -0.101190469 | 6.657124108 | 0.922379109 |
| vegetation index (MSAVI) during all seasons (100m buffer) at age 14 | -0.04 [-0.25, 0.17] | 0.094865489 | -0.449958219 | 11.33717525 | 0.661222742 |
| vegetation index (MSAVI) during all seasons (500m buffer) at age 14 | -0.05 [-0.12, 0.03] | 0.038825373 | -1.174778663 | 166.7059517 | 0.241758585 |
| vegetation index (NDVI) during greenest season (100m buffer) at age 14 | 0.01 [-0.21, 0.23] | 0.109092805 | 0.105694233 | 73.71195524 | 0.916112059 |
| treecover (500m buffer) at age 14 | 0.02 [-0.02, 0.06] | 0.020337161 | 1.215710856 | 746.4221354 | 0.224479439 |

### Twin Modelling

#### eTable 11 - Twin modelling assumption tests: Equality of Means and Variances for the univariate Twin model on unadjusted (full) EA

eTable 11: Tests for equality of means were conducted using paired t-tests (within-pair) and unpaired t-tests (between zygosity; DZ=dizygotic, MZ=monozygotic). Opposite-sex DZ twins were excluded from the Twin models. Tests for equality of variances were conducted using F-tests. Statistical significance ($p < .05$) indicates a violation of the equal environments assumption or sampling bias required for standard twin modelling.

| Outcome | Comparison | Test Statistic (t) | p-value | Assumption met |
|---|---|---|---|---|
| PCGrimAge | Mean: MZ Twin1 vs Twin2 | -1.7 | 0.091 | yes |
| PCGrimAge | Mean: DZs Twin1 vs Twin2 | 0.07 | 0.946 | yes |
| PCGrimAge | Mean: MZ Twin1 vs DZs Twin1 | -0.15 | 0.881 | yes |
| PCGrimAge | Variance: MZ Twin1 vs Twin2 | 0.94 | 0.688 | yes |
| PCGrimAge | Variance: DZs Twin1 vs Twin2 | 0.99 | 0.963 | yes |
| PCGrimAge | Variance: MZ Twin1 vs DZs Twin1 | 0.95 | 0.754 | yes |
| DunedinPACE | Mean: MZ Twin1 vs Twin2 | -0.1 | 0.92 | yes |
| DunedinPACE | Mean: DZs Twin1 vs Twin2 | 0.08 | 0.933 | yes |
| DunedinPACE | Mean: MZ Twin1 vs DZs Twin1 | -0.88 | 0.38 | yes |
| DunedinPACE | Variance: MZ Twin1 vs Twin2 | 0.95 | 0.757 | yes |
| DunedinPACE | Variance: DZs Twin1 vs Twin2 | 0.84 | 0.328 | yes |
| DunedinPACE | Variance: MZ Twin1 vs DZs Twin1 | 1.19 | 0.318 | yes |

#### eTable 12 - Twin modelling assumption tests: Equality of Means and Variances for the univariate Twin model on residual (unexplained) EA

eTable 12: Tests for equality of means were conducted using paired t-tests (within-pair) and unpaired t-tests (between zygosity; DZ=dizygotic, MZ=monozygotic). Opposite-sex DZ twins were excluded from the Twin models. Tests for equality of variances were conducted using F-tests. Statistical significance ($p < .05$) indicates a violation of the equal environments assumption or sampling bias required for standard twin modelling.

| Outcome | Comparison | Test Statistic (t) | p-value | Assumption met |
|---|---|---|---|---|
| PCGrimAge | Mean: MZ Twin1 vs Twin2 | -1.31 | 0.196 | yes |
| PCGrimAge | Mean: DZs Twin1 vs Twin2 | -0.03 | 0.952 | yes |
| PCGrimAge | Mean: MZ Twin1 vs DZs Twin1 | 0.14 | 0.873 | yes |
| PCGrimAge | Variance: MZ Twin1 vs Twin2 | 1.03 | 0.828 | yes |
| PCGrimAge | Variance: DZs Twin1 vs Twin2 | 0.96 | 0.811 | yes |
| PCGrimAge | Variance: MZ Twin1 vs DZs Twin1 | 0.93 | 0.637 | yes |
| DunedinPACE | Mean: MZ Twin1 vs Twin2 | -0.04 | 0.893 | yes |
| DunedinPACE | Mean: DZs Twin1 vs Twin2 | 0.13 | 0.889 | yes |
| DunedinPACE | Mean: MZ Twin1 vs DZs Twin1 | -0.11 | 0.936 | yes |
| DunedinPACE | Variance: MZ Twin1 vs Twin2 | 1.01 | 0.954 | yes |
| DunedinPACE | Variance: DZs Twin1 vs Twin2 | 0.85 | 0.326 | yes |
| DunedinPACE | Variance: MZ Twin1 vs DZs Twin1 | 0.98 | 0.872 | yes |

#### eTable 13 - Descriptive Statistics and Shapiro-Wilk Tests for Normality of unadjusted (full) EA

eTable 13: Results of the Shapiro-Wilk test for normal distribution of unadjusted (full) EA. Although significant deviations were detected, skewness and kurtosis values remained within ranges (e.g., skewness < 2.0) typically considered acceptable for Maximum Likelihood estimation in twin modelling.

| Outcome | Group | N | Shapiro-Wilk W | p-value | Skewness | Kurtosis | Normal distribution |
|---|---|---|---|---|---|---|---|
| GrimAge | MZ Twin 1 | 164 | 0.976 | **0.007** | 0.39 | 1.73 | no |
| GrimAge | MZ Twin 2 | 164 | 0.94 | **< .001** | 0.97 | 2.95 | no |
| GrimAge | DZ Twin 1 | 122 | 0.968 | **0.005** | 0.57 | 0 | no |
| GrimAge | DZ Twin 2 | 122 | 0.971 | **0.01** | 0.64 | 0.41 | no |
| DunedinPACE | MZ Twin 1 | 164 | 0.978 | **0.012** | 0.55 | 0.47 | no |
| DunedinPACE | MZ Twin 2 | 164 | 0.982 | **0.029** | 0.51 | 0.42 | no |
| DunedinPACE | DZ Twin 1 | 122 | 0.993 | 0.811 | 0.21 | -0.01 | yes |
| DunedinPACE | DZ Twin 2 | 122 | 0.97 | **0.008** | 0.64 | 0.69 | no |

*Note.* MZ = Monozygotic; DZ = Dizygotic; *N* = Number of individuals.

#### eFigure 6 - Tests for Normality of unadjusted (full) EA

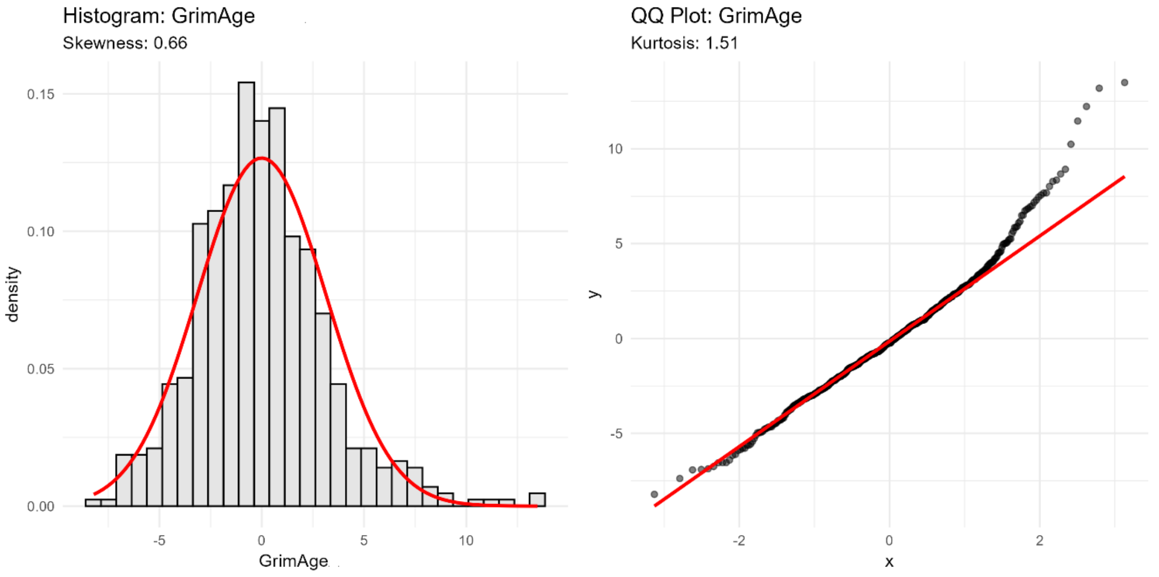

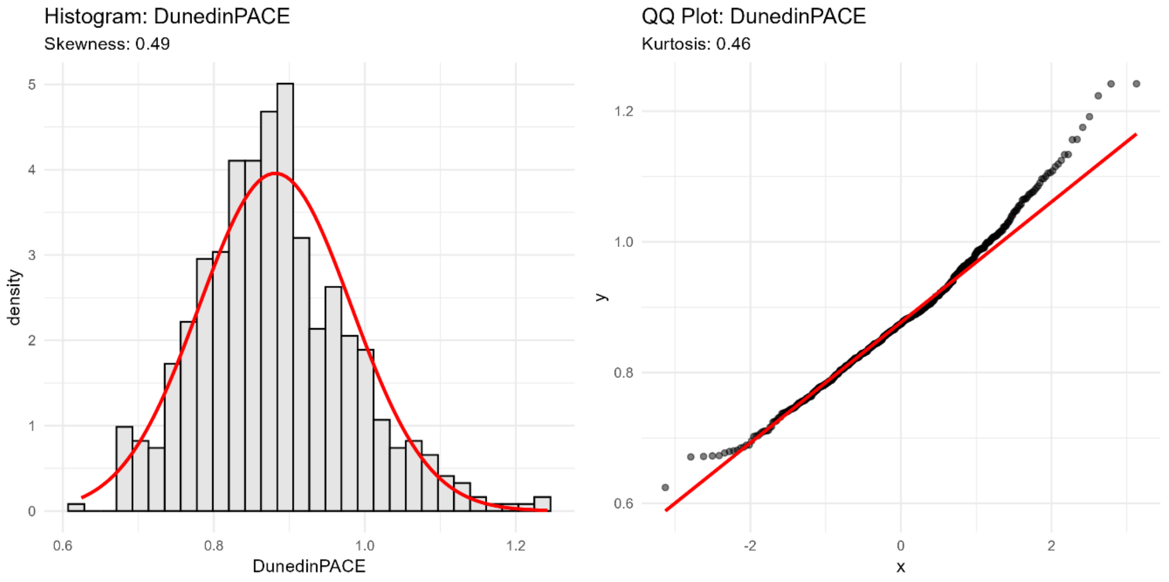

eFigure 6 – Histograms of unadjusted (full) $EA_{GrimAge}$ and $EA_{DunedinPACE}$ in the final analytical sample excluding opposite se DZ twins (N=572). Visual inspection of histograms and QQ-plots confirmed the distributions were sufficiently normal for Maximum Likelihood estimation; therefore, raw data were used to preserve biological interpretability.

#### eTable 14 - Descriptive Statistics and Shapiro-Wilk Tests for Normality of residual (unexplained) EA

eTable 14: Results of the Shapiro-Wilk test for normal distribution of residual (unexplained) EA. Although significant deviations were detected, skewness and kurtosis values remained within ranges (e.g., skewness < 2.0) typically considered acceptable for Maximum Likelihood estimation in twin modelling.

| Outcome | Group | N | Shapiro-Wilk W | p-value | Skewness | Kurtosis | Normal distribution |
|---|---|---|---|---|---|---|---|
| GrimAge | MZ Twin 1 | 164 | 0.955 | **< .001** | 0.86 | 3.83 | no |
| GrimAge | MZ Twin 2 | 164 | 0.922 | **< .001** | 1.25 | 4.04 | no |
| GrimAge | DZ Twin 1 | 122 | 0.992 | 0.304 | 0.11 | 0.21 | yes |
| GrimAge | DZ Twin 2 | 122 | 0.916 | **< .001** | 1.44 | 4.66 | no |
| DunedinPACE | MZ Twin 1 | 164 | 0.985 | 0.085 | 0.48 | 0.55 | yes |
| DunedinPACE | MZ Twin 2 | 164 | 0.982 | **0.029** | 0.43 | 1.02 | no |
| DunedinPACE | DZ Twin 1 | 122 | 0.989 | 0.108 | 0.3 | 0.49 | yes |
| DunedinPACE | DZ Twin 2 | 122 | 0.943 | **< .001** | 0.91 | 4.18 | no |

#### eFigure 7- Tests for Normality of residual (unexplained) EA

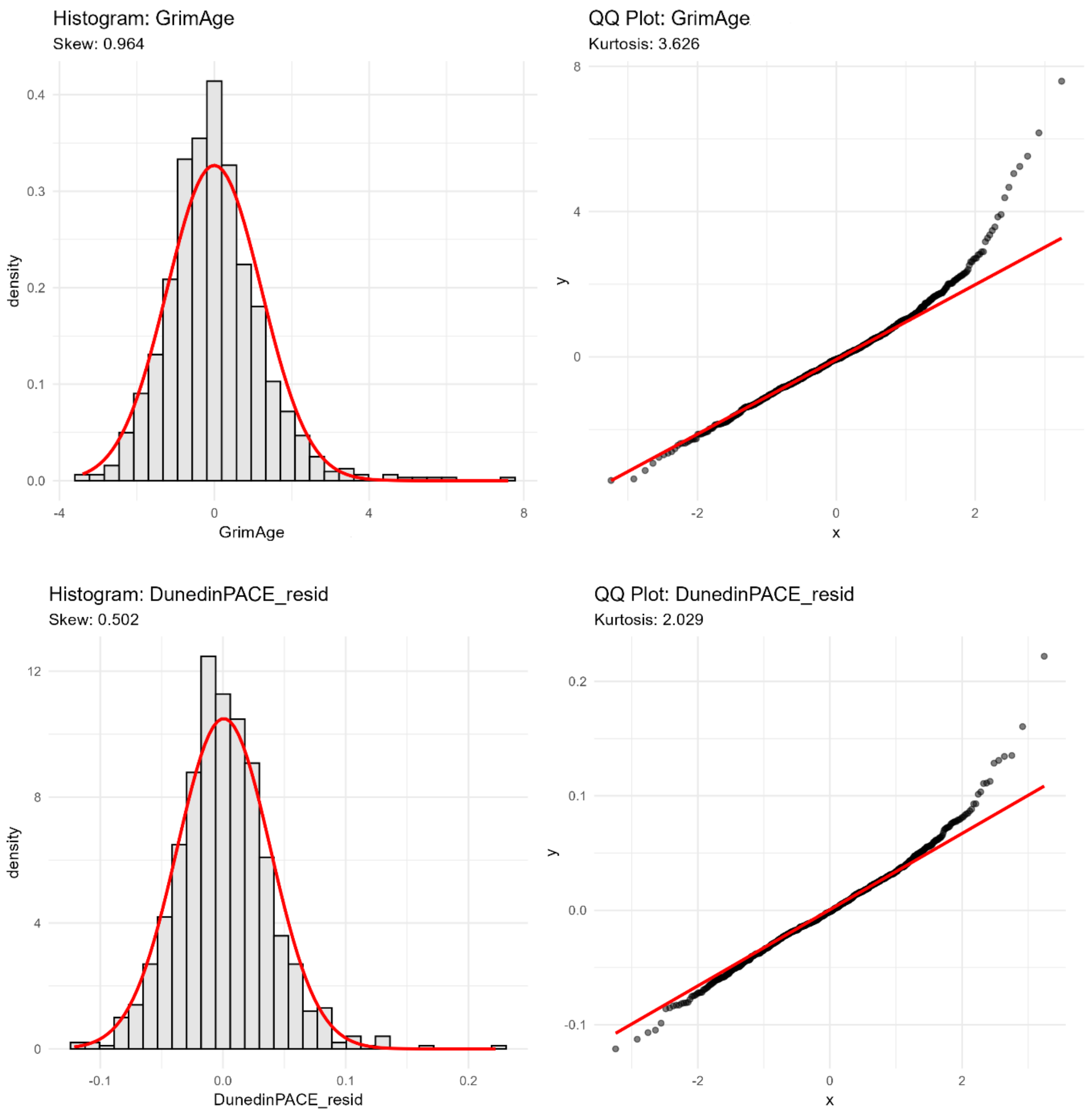

eFigure 7 – Histograms of residual (unexplained) $EA_{GrimAge}$ and $EA_{DunedinPACE}$ in the final analytical sample excluding opposite se DZ twins (N=572). Visual inspection of histograms and QQ-plots confirmed the distributions were sufficiently normal for Maximum Likelihood estimation; therefore, raw data were used to preserve biological interpretability.

#### eTable 15 - Univariate Twin Model Fit statistics and variance decomposition for full (unadjusted) variance

eTable 15 presents the fit indices and standardized variance components for Additive Genetics (A), Shared Environment (C), Dominance Genetics (D), and Unique Environment (E). Values in parentheses represent the 95% Wald confidence intervals. Model fit was evaluated using Minus 2 Log-Likelihood (-2LL), Akaike Information Criterion (AIC), and Root Mean Square Error of Approximation (RMSEA). The AE submodel was compared against the best-fitting full model (ADE) using a Chi-squared ($X^2$) difference test.

| Outcome | Model | Median -2LL | Median df | AIC | RMSEA | Comparison | ΔX² (p-value) | a² [95% CI] | c² [95% CI] | d² [95% CI] | e² [95% CI] |
|---|---|---|---|---|---|---|---|---|---|---|---|
| GrimAge | ACE | 2122.69 | 752 | 618.69 | 0.091 | saturated | — | 0.38 [0.16;0.47] | 0.00 [0.00;0.00] | — | 0.58 [0.54;0.81] |
| GrimAge | ADE | 2114.45 | 752 | 610.45 | 0.068 | — | — | 0 | — | 0.45 [0.27;58] | 0.55 [0.44;0.70] |
| GrimAge | AE | 2122.69 | 753 | 616.69 | 0.082 | vs. ACE | 8.24 (0.004) | 0.38 [0.17;0.48] | — | — | 0.62 [0.54;0.81] |
| DunedinPACE | ACE | 2144.5 | 752 | 640.5 | 0.07 | saturated | — | 0.16 [0.00;0.31] | 0.00 [0.00;0.00] | — | 0.84 [0.68;1.00] |
| DunedinPACE | ADE | 2141.14 | 752 | 637.14 | 0.058 | — | — | 0 | — | 0.24 [0.06;0.42] | 0.76 [0.58;0.93] |
| DunedinPACE | AE | 2144.5 | 753 | 638.5 | 0.061 | vs. ACE | 3.36 (0.067) | 0.16 [0.01;0.31] | — | — | 0.84 [0.68;1.00] |

#### eTable 16 - Univariate Twin Model Fit statistics and variance decomposition for residual (unexplained) variance

eTable 16 presents the fit indices and standardized variance components for Additive Genetics (A), Shared Environment (C), Dominance Genetics (D), and Unique Environment (E). Values in parentheses represent the 95% Wald confidence intervals. Model fit was evaluated using Minus 2 Log-Likelihood (-2LL), Akaike Information Criterion (AIC), and Root Mean Square Error of Approximation (RMSEA). The AE submodel was compared against the best-fitting full model (ADE) using a Chi-squared ($X^2$) difference test.

| Outcome | Model | Median -2LL | Median df | AIC | RMSEA | Comparison | ΔX² (p-value) | a² [95% CI] | c² [95% CI] | d² [95% CI] | e² [95% CI] |
|---|---|---|---|---|---|---|---|---|---|---|---|
| GrimAge | ACE | 2720.28 | 842 | 1036.28 | 0.086 | saturated | — | 0.38 [0.25;0.51] | 0 [0.00;0.00] | — | 0.58 [0.54;0.81] |
| GrimAge | ADE | 2712.55 | 842 | 1028.55 | 0.066 | vs. ACE | — | 0 | — | 0.45 [0.27;58] | 0.55 [0.44;0.70] |
| GrimAge | AE | 2720.28 | 843 | 1034.28 | 0.076 | vs. ACE | 7.684 (0.004) | 0.38 [0.17;0.48] | — | — | 0.62 [0.54;0.81] |
| DunedinPACE | ACE | -3125.63 | 842 | -4809.63 | 0.155 | saturated | — | 0.16 [0.00;0.31] | 0.00 [0.00;0.00] | — | 0.84 [0.68;1.00] |
| DunedinPACE | ADE | -3125.63 | 842 | -4809.63 | 0.155 | vs. ACE | — | 0 | — | 0.24 [0.06;0.42] | 0.76 [0.58;0.93] |
| DunedinPACE | AE | -3125.63 | 843 | -4811.63 | 0.14 | vs. ACE | 2.929 (0.087) | 0.16 [0.01;0.31] | — | — | 0.84 [0.68;1.00] |

### Follow-up analyses

#### eFigure 8 – Association between green space size and biological aging (DunedinPACE) stratified by land use type.

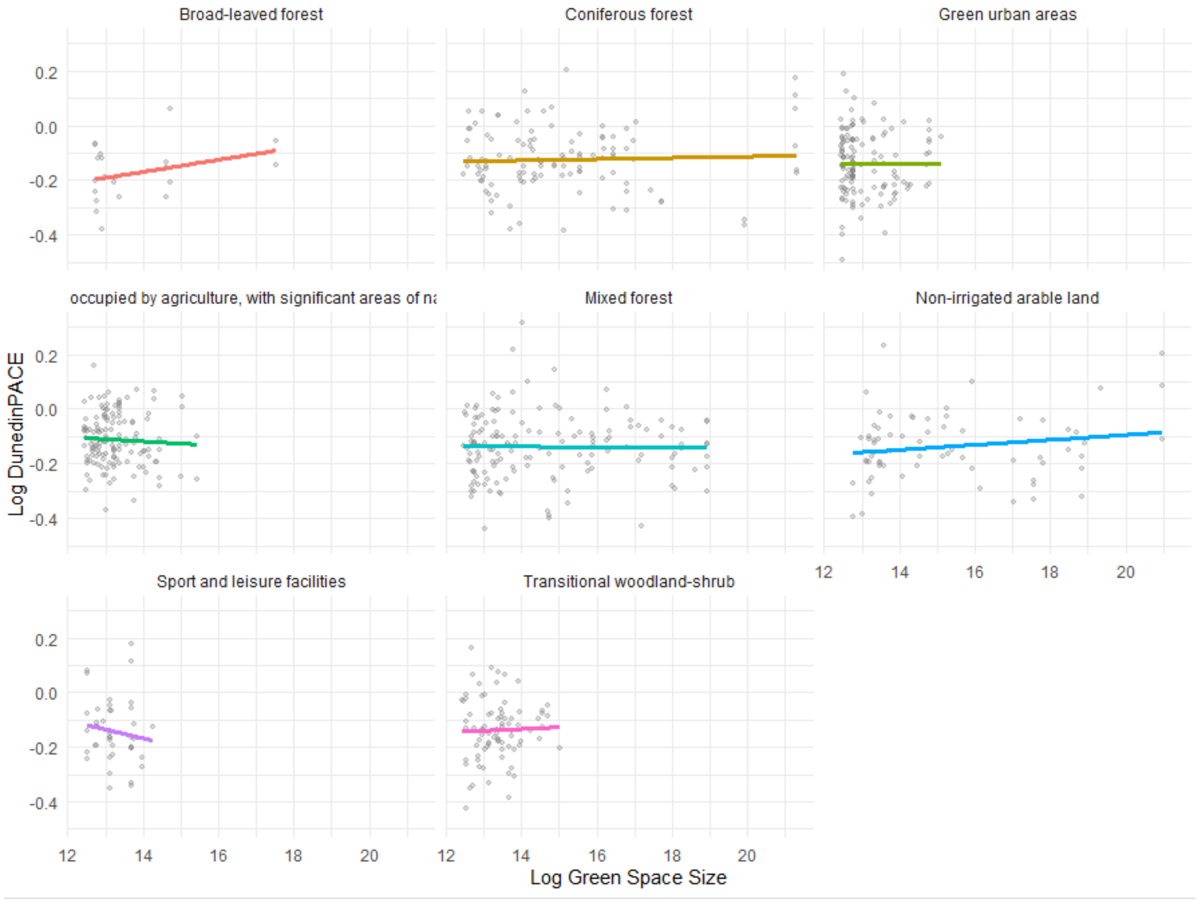

eFigure 8: Scatter plots display the unadjusted relationship between the log-transformed size of green spaces and the log-transformed pace of aging (DunedinPACE). Panels represent eight distinct land types: broad-leaved forest, coniferous forest, green urban areas, agricultural land with significant natural vegetation, mixed forest, non-irrigated arable land, sport and leisure facilities, and transitional woodland-shrub. Solid colored lines indicate the raw linear trend for each land type. Points represent individual observations.

#### eTable 17 – Adjusted linear regression slopes for the association between log-transformed green space size and DunedinPACE across distinct land-use categories.

eTable 17 shows the estimates representing the change in log-DunedinPACE per unit increase in log-green space size, adjusted for covariates.

| **Land Type** | **N** | **Slope (95% CI)** | **p-value** |
|---|---|---|---|
| Broad-leaved forest | 21 | 0.016 [-0.017, 0.049] | 0.336 |
| Coniferous forest | 111 | 0.003 [-0.007, 0.012] | 0.556 |
| Green urban areas | 133 | 0.004 [-0.023, 0.03] | 0.777 |
| Land principally occupied by agriculture, with significant areas of natural vegetation | 136 | -0.009 [-0.037, 0.02] | 0.546 |
| Mixed forest | 147 | 0 [-0.009, 0.009] | 0.946 |
| Non-irrigated arable land | 69 | 0.007 [-0.004, 0.018] | 0.218 |
| Sport and leisure facilities | 44 | -0.035 [-0.105, 0.034] | 0.321 |
| Transitional woodland-shrub | 81 | 0.013 [-0.026, 0.052] | 0.52 |

#### eTable 18 – Regression analysis of landscape type on aerial black carbon

eTable 18 presents results from a multivariate regression model evaluating the impact of landscape type (“Urban” vs. “Farming”) on aerial black carbon content. The model was adjusted for covariates.

| | estimate [95%CI] | p-value |
|---|---|---|
| baseline ("Farming", MZ, female sex) | 0.125 [-1.259, 1.508] | 0.859 |
| landscape type "Urban" vs. "Farming" | 0.25 [0.165, 0.0335] | <0.001 |
| *covariates* | | |
| chronological age | 0 [-0.06, 0.06] | 0.993 |
| male sex vs. female sex | 0.017 [-0.051, 0.085] | 0.62 |
| DZ vs. MZ | -0.035 [-0.134, 0.064] | 0.486 |
| OSDZ vs. MZ | 0.032 [-0.076, 0.139] | 0.561 |
| maternal age at birth | 0.008 [-0.002, 0.017] | 0.107 |
| paternal education score | 0.014 [-0.006, 0.034] | 0.18 |
| maternal education score | 0.004 [-0.019, 0.026] | 0.753 |

#### eFigure 9 - Distribution of black carbon exposure levels at age 22 stratified by land type

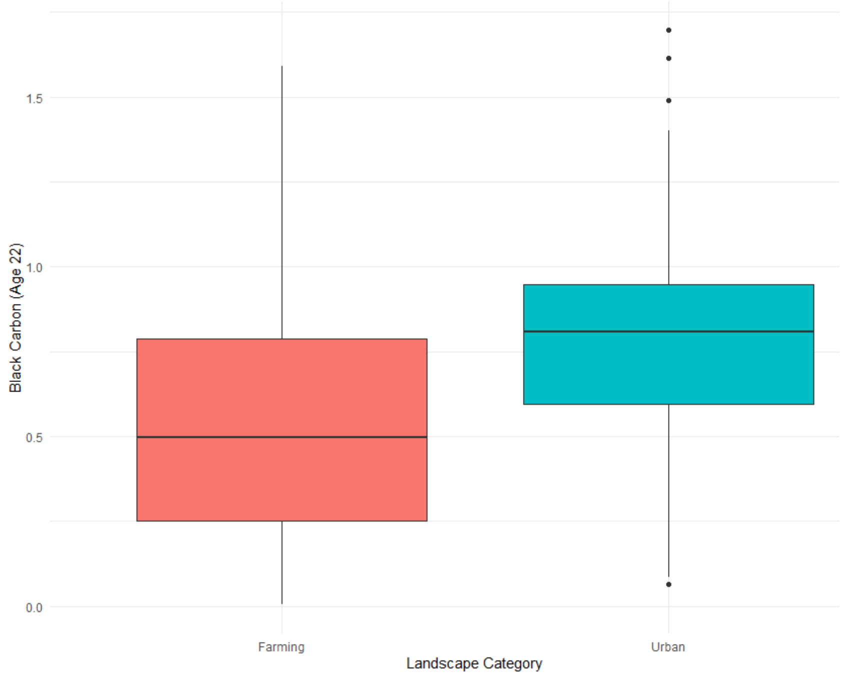

eFigure 9 displays the results of the Linear Mixed Model performed on aerial black carbon concentrations between residents of "Farming" environments (aggregating agricultural and non-irrigated arable land) versus "Urban" green spaces (aggregating sport/leisure facilities and green urban areas). The horizontal bar represents the median; the box encloses the interquartile range. The "Urban" category demonstrates a higher median black carbon level compared to the "Farming" category.
